# Macro-parasite transmission in dynamic seasonal environment: Basic Reproductive Number, endemicity, and control

**DOI:** 10.1101/19012245

**Authors:** Q. Huang, D. Gurarie, M. Ndeffo-Mbah, E. Li, CH. King

## Abstract

Seasonality of transmission environment, which includes snail populations and habitats, or human-snail contact patterns, can affect the dynamics of schistosomiasis infection, and control outcomes. Conventional modeling approaches often ignore or oversimplify it by applying ‘seasonal mean’ formulation. Mathematically, such ‘averaging’ is justified when model outputs/quantities of interest depend linearly on input variables. That is not generally the case for macroparasite transmission models, where model outputs are nonlinear functions of seasonality fashion.

Another commonly used approach for Schistosomiasis modeling is a reduction of coupled human-snail system to a single ‘human equation’, via quasi-stationary snail (intermediate host) dynamics. The basic questions arising from these approaches are whether such ‘seasonal averaging’ and ‘intermediate host reduction’ are suitable for highly variable/seasonal environments, and what implications these methods have on models’ predictive potential of control interventions.

Here we address these questions by using a combination of mathematical analysis and numerical simulation of two commonly used models for macroparasite transmission, MacDonald (MWB), and stratified worm burden (SWB) snail-human systems. We showed that predictions from ‘seasonal averaging’ models can depart significantly from those of quasi-stationary models. Typically, seasonality would lower endemicity and sustained infection, vs. stationary system with comparable transmission inputs. Furthermore, discrepancies between the two models (‘seasonal’ and its ‘stationary mean’) increase with amplitude (or variance) of seasonality. So sufficiently high variability can render infection unsustainable. Similar discrepancies were observed between coupled and reduced ‘single host’ models, with reduced model overpredicting sustained endemicity. Seasonal variability of transmission raises the question of optimal control timing. Using dynamic simulation, we show that optimal timing of repeated MDA is about half season past the snail peak, where snail population attains its minimal value. Compared to sub-optimal timing, such strategy can reduce human worm burden by factor 2 after 5-6 rounds of MDA. We also extended our models for dynamic snail populations, which allowed us to study the effect of repeated molluscicide, or combined strategy (MDA + molluscicide). The optimal time for molluscicide alone is the end or the start of season, and combined strategy can give additional reduction, and in some cases lead to elimination.

Overall, reduced sustainability in seasonal environment makes it more amenable to control interventions, compared to stationary environment.

## Introduction

Seasonal transmission of an infectious disease is a common phenomenon, defined by time dependent transmission rates that vary over the season. Schistosomiasis is a case example where the seasonality is related to snail populations linked to weather patterns (rainfall, temperature), and/or behavioral factors (human-snail contacts). But mathematical models of Schistosome transmission rarely account for seasonality, with implication to accuracy of model prediction and opportunity to gain insight into public health practice, like optimal timing of interventions.

Basic reproduction number (*R*_0_) is a fundamental concept widely used in population biology and infectious disease modeling. In the context of infectious diseases, *R*_0_ is used to assess the magnitude of disease outbreak, spread or endemic (equilibrium) level [^1^]. Its applications range from modeling of human-to-human communicable diseases to vector-mediated systems for macroparasite diseases, such as Ross-MacDonald models of schistosomiasis (see e.g. ^2, 3, 4^,^5^,^6^). Though *R*_0_ was first explicitly derived for simple single population models, this concept was further extended to more complex meta-population systems, e.g. spatially connected environment with multiple host sites (see e.g. ^7, 8,9, 10,11^).

Mathematically, *R*_0_ is most appropriate for stationary environment where disease transmission parameters are kept fixed. However, in many cases, such environment is highly variable, e.g. seasonal change of vector population or contact patterns, as well as abrupt changes caused by control interventions.

One way to incorporate such variability in mathematical models is through appropriate ‘seasonal averaging’ of environment and/or behavioral inputs. However, little effort was spent to assess the effect of ‘seasonal averaging’ for analysis of disease predictions, and control outcomes. In general, the use of averaging methods in non-stationary dynamical systems can be justified when variability is marginal relative to baseline mean state or when quantities of interest (e.g. dynamic variables or outputs) depend linearly on variable inputs. Under such conditions, the ‘averaged’ stationary model with properly adjusted coefficients is able to reproduce approximate ‘mean’ behavior of the non-stationary (time periodic) system. However, neither of these conditions holds for nonlinear disease transmission models in variable seasonal environment. The outputs of such system are nonlinear functions of its input variables, and they can depart significantly from the expected ‘mean’ behavior.

Besides seasonal averaging, another commonly used approximation in modeling vector-host transmission is a reduction of the coupled system to a single host equation, via quasi-equilibration of fast vector variables. Such reduction is commonly justified by distinct time scales: slow ‘host-parasite dynamics’ vs. fast ‘vector-intermediate host’. Indeed, in the context of schistosomiasis time scales for snail dynamics vary from weeks-to-months, while human and worm scales are order of magnitude slower (years).

Reduced models are convenient for theoretical analysis and for numeric simulations (see e.g. ^2^, ^3^). Such reduction procedures however, require more careful analysis beyond simple ‘time-scale’ heuristics, to assess possible departure of the reduced or averaged models (‘single-host’ or ‘seasonal-mean’) from their fully coupled, non-stationary precursor.

Here we assess the effect of seasonal averaging and intermediate host reduction for schistosomiasis transmission in seasonal environment, and their implications on predicting control outcomes (MDA). On human side, we shall mostly use mean worm burden (MWB) MacDonald formulation (^4, 5-7^), but make a few comment on other possible approaches, like stratified worm burden system (^8-10^). Seasonal transmission environment can appear as variable snail population and/or variable human-snail contacts.

In this paper, we mainly focus on seasonal snail variability (their population density) that determines their infection level, and snail-to-human transmission rate. Mathematically, different functions can be used to account for variable snail population, under different environmental conditions(precipitation, temperature, et al) (see e.g. ^11,12, 13, 14, 15, 16^). The two commonly observed patterns include: (i) moderate snail variability about ‘mean value’ with approximately equal high and low seasons, (ii) long dry season (with low / zero snail density and transmission) followed by short rainy season, with bursting snail population and intensified transmission rate.

We shall employ two mathematical functions for such seasonal patterns, namely (i) trigonometric function 1+ *a* cos (2*πt*) with amplitude 0 < *a* < 1, for alternating high/low seasons of equal duration, (ii) concentrated peak density over low-level background with prescribed seasonal mean value =1. The latter also have a version of amplitude parameter (0 < *a* < 1), but its meaning and implication are different.

Earlier theoretical papers (^17^,^18^) studied seasonally varying (periodic) human-snail contact patterns and their impact on schistosomiasis endemic state and persistence. In particular, ^18^ estimated endemicity thresholds for periodically varying MacDonald system. Here we change the focus from ‘seasonal contact’ to ‘seasonal snail population’, and extend the scope of their analysis by exploring the implications of seasonality for sustained infection, and control interventions, MDA and molluscicide.

The bulk of our analysis employs prescribed snail population (density) function *N* (*t*). The study of molluscicide control however, requires a dynamic snail population that could respond to an abrupt change, environmental and/or human-made interventions. Such resource-driven models of snail population biology were developed in several papers (e.g. ^15, 16^, ^19^). These works however, did not apply dynamic snail models for transmission/control analysis. Our paper attempts to fill this gap. Alongside prescribed function *N* (*t*) we develop model of resource driven seasonal snail, and apply it to study molluscicide.

Two basic parameters employed in our analysis are basic reproduction number *R*_0_ (of ‘mean’ stationary system), and amplitude (*a*) of seasonal variability.

By combining mathematical analysis with numeric simulations, we address several questions:

i. combined effect of (*R*_0_, *a*) on periodic/stationary patterns of human-snail infection and their seasonal means. Among other results we identified parameter ranges where transmission becomes unsustainable for different types of seasonality.
ii. We explore long terms effect of repeated MDA in different model formulations. Once again, ‘reduced’ and ‘seasonal-mean’ MacDonald system can depart significantly from compete (fully coupled) dynamical system.

We apply our model to study optimal seasonal timing of repeated interventions MDA, molluscicide, or integrated strategies, that would achieve maximal parasite burden reduction over limited time-span (6-10 years). For MDA alone, we estimated optimal seasonal timing in the range ¼ - ½ season past snail peak. Optimal effect of molluscicide on worm reduction is achieved at the end or start of season (preceding snail peak). For mixed strategy (MDA + molluscicide) we can use estimated optimal timing of each intervention, to achieve best results.

## Modeling setup

### Stationary transmission environment: *R*_0_ analysis

**Simple MacDonald** system for 2 variables *w*(*t*)-human MWB, and *y* (*t*) - infected snail prevalence, consists of coupled differential equations

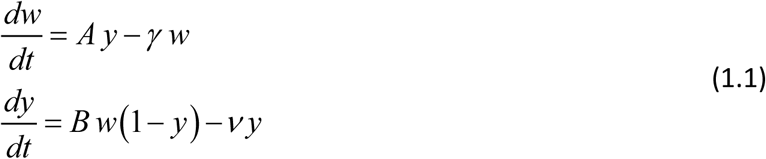

with transmission coefficients *A, B*, and worm/snail loss rates (*γ, ν*). The former *A* is proportional to snail density *N*, the latter *B* depends on human population size *H*. System (1.1) can be rescaled in dimensionless form, given by a single parameter 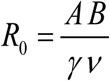 (see Box 1),

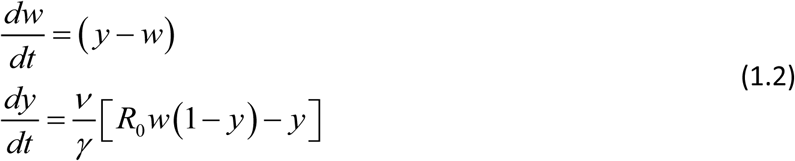

Parameter *R* _0_ is responsible for endemic equilibria of (1.2) and for endemic to zero transition. A simple illustration is herd immunity, vaccine coverage fraction (*f* > 1−1/ *R*_0_) to prevent an outbreak.

Second order system (1.2) can be reduced by replacing ‘fast’ snail equation is with its quasi-equilibrium value (see Box 1) due to’ short’ snail time scales (months) compared to ‘slow’ human-worm dynamics (years). The reduced MacDonald system is a single equation

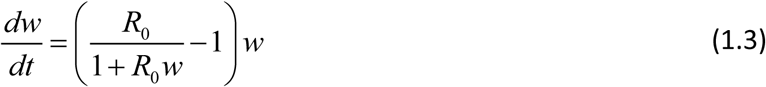

Both full and reduced systems (1.2)- (1.3) share endemic equilibrium in a stationary environment. Significant differences however, could arise is seasonal environment.

#### Box 1

Rescaled (dimensionless) MacDonald system (1.1) for variable 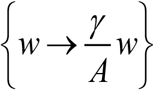, 0<(*w,y*)<1 depends on a single parameter 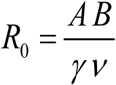. It consists of human and snail equations: 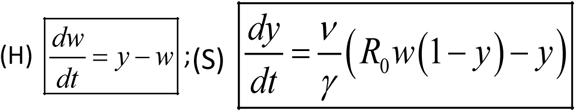. In stationary environment such (H-S) system has stable endemic equilibrium, *w*^*^ = *y*^*^ = 1−1/ *R*, provided *R*_0_ > 1, and unstable infection-free (zero) state. The (S) equation can be further reduced to its quasi-equilibrium state, 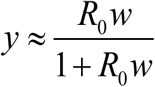, assuming snail turnover rate compared to worm mortality (*γ* /*ν* ≫1). Then (H-S) system becomes a single worm equation (W) 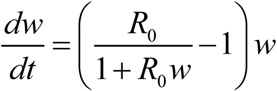. Systems (H-S) and (W) have identical equilibria, and their solutions remain close under stationary or slowly varying environmental change (e.g. near constant snail density and exposure/ contamination rates).

#### MacDonald system with mating and aggregation

Many conventional approaches, going back to the original MacDonald paper ^4^, assume worm burden is distributed in host population according to negative binomial (NB) pattern with mean value ***w*(*t*)**, and aggregation parameter *k* > 0 (^5,7,20,21^). In such setting, the force of snail infection (= *Bw*) of (1.1) is no more proportional to MWB ***w*(*t*)**. Instead, one should use mean mated count (MCB), which depends on worm distribution in host community. For assumed NB – distribution, it can be estimated via so-called mating function 0 < *ϕ*_*k*_ (*w*) < 1, namely MCB = *w ϕ*_*k*_ (*w*). Equations (1.1) are modified accordingly

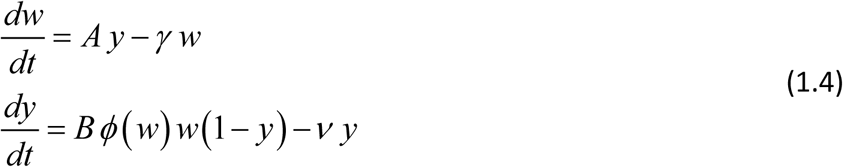

After rescaling (Box 1) we recast it as

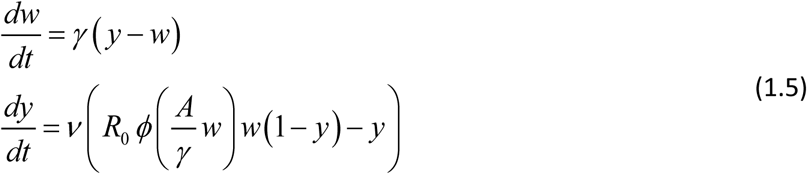

or its reduced (single-host) version

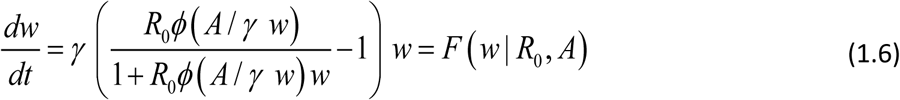

Compared to (1.2) modified system (1.5) depends on 2 dimensionless parameters, transmission coefficients (*A* / *γ* ; *B* /*ν*), or pair 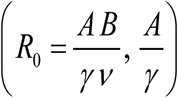, rather than single *R*_0_. There are other fundamental differences between simple MacDonald (1.2), and its modified version (1.5). The former has two equilibria (zero-endemic), whose stability types are determined by *R*_0_ (*R*_0_ > 1 - stable endemic, *R*_0_ <1-stable zero), *saddle-no*de type. Modified system (1.5) is *bistable* for sufficiently large *R*_0_ > *R*_*c*_ > 1, above critical value *R*_*c*_. Figure F of Supplement illustrates critical *R*_*c*_ = 2.4, for specific choice of transmission coefficients. It has stable equilibria (zero + endemic), and intermediate (unstable) breakpoint (see Supplement, Figure Mated MacDonald). Such breakpoints have important implications for MDA control, explained below.

### Seasonal environment

#### Non-stationary

transmission dynamics can arise from multiple sources, including natural (seasonal snail populations), behavioral (human-snail exposure/contamination), or repeated interventions, like MDA or molluscicide. In this section we focus on variable snail population, given by function *N*(*t*) (see e.g. ^19, 15^). There are different types of snail environment, that make time dependent snail population *N* (*t*). We do not attempt to classify and study all such patterns of natural variability, but confine our analysis to 2 typical periodic functions

I. Trigonometric: *N* (*t, a*) = 1+ *a* cos (2*π t*) of amplitude 0 ≤ *a* ≤ 1, whereby season is evenly divided between high / low snail density (above and below-average).
II. Peak-type *N* (*t, a*) of amplitude 0 ≤ *a* < 1 (Figure *1*), defined via elliptic theta function. Amplitude *a* has different meaning now. Large *a* (close to 1) features sharp seasonal peak of short duration about *t=0*, followed by long dry period (*N* (*t*) ≈ 0) over the remaining ‘dry’ season.

Data from different studies on snail abundance support such patterns, e.g. (^13, 19 22^). In both cases, *N* (*t, a*) is rescaled so that its seasonal mean, 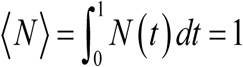. Function *N* (*t, a*) represents relative snail density, its absolute ‘mean’ values ⟨*N*⟩ is included into parameter *R*_0_.

The variability of function *N* (*t, a*) can be measured by its variance, *σ*^2^ = ⟨*N*^2^⟩ −1. In case (i), 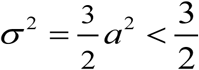 is limited, while case (ii) allows unlimited values of variance, as (*a* → 1) and population peak sharpness.

System (1.7) with periodic *N* (*t*) of either type has stable periodic solutions {*w*^*^ (*t*), *y*^*^ (*t*)} determined by (*R*_0_, *a*), which play the role of endemic equilibria for stationary model.

##### Box 2

Periodic seasonal forcing is determined by relative snail population density *N* (*t*) with mean 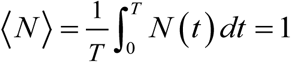, either trigonometric function *N* (*t, a*) = 1+ *a* cos (2*π t*) of amplitude (0 ≤ *a* < 1), or ‘single-peak’ type *N* (*t*) (Figure 1). Some modifications are needed to accommodate variable snail population, namely

i. prevalence variable *y* (*t*) in (1.1) is replaced by infected snail density (0 < *y* (*t*) < *N* (*t*)), and susceptible pool is given by clipped function **[*N* (*t*) − *y* (*t*)]**_**+**_
ii. fixed snail mortality (= rescaled rate 1 of (1.2)) is replaced by time dependent rate function *μ* (*t*) = max (1; − *N*′(*t*) /*ν N* (*t*)).

Such dynamic (time-dependent) function *μ* (*t*) should replace its rescaled ‘natural mortality’ (= 1) in case of steep population drop (−*N*′(*t*) / *N* (*t*) >*ν*), lest function *y* (*t*) (infected snail pool) would exceeds total population *N* (*t*). In our setup, natural snail mortality (*ν*) refers to abundant food supply, while steep population decline (e.g. dry season), means increased mortality max [*ν*, −*N*′(*t*) / *N* (*t*)], hence function *μ* (*t*). The reduced worm equation (1.3) is modified accordingly (equation (1.8)). In either case, full (2D) system (1.7) or reduced (1.8), there exists a stable periodic solution *w*^*^ (*t*), which plays the role of stable endemic equilibrium. In can be computed from the fixed point of the corresponding Poincare (period) map.

**Figure 1:**
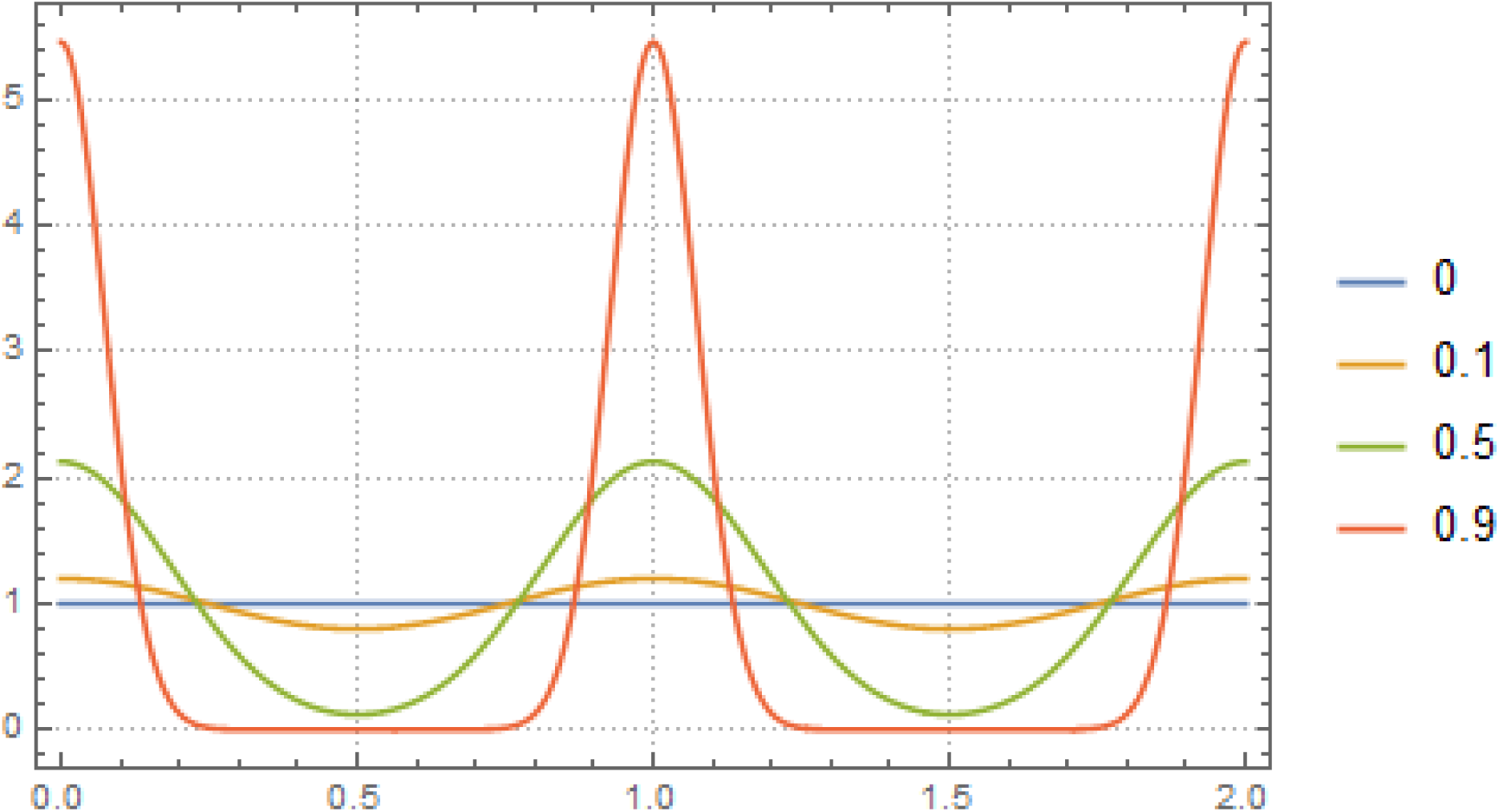
Examples of type II ‘peak seasonality’, modeled by elliptic theta function, 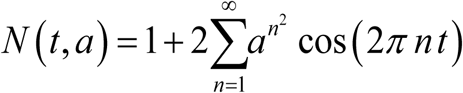 of amplitude a. Functions *N* (*t, a*) are normalized, so that their seasonal mean 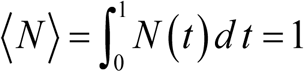.

To derive ‘seasonal’ dynamic equations, similar to (1.2)-(1.3), we can follow the scheme of Box 1, but some modifications are needed to account for non-stationary ***N* (*t*)**. Specifically, infected prevalence variable 0 < *y* (*t*) < 1 in (1.2) should be replaced by infected snail density (0 < *y* (*t*) < *N* (*t*)), and constant snail mortality ***ν*** by time-dependent function *μ* (*t*) (Box 2). The resulting modified MacDonald takes the form

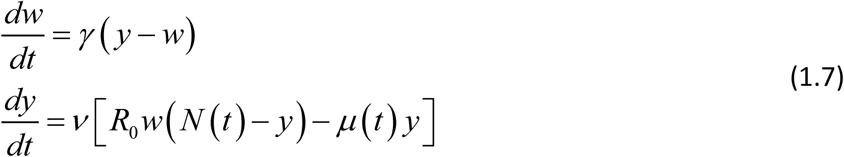

Its reduced version is given by

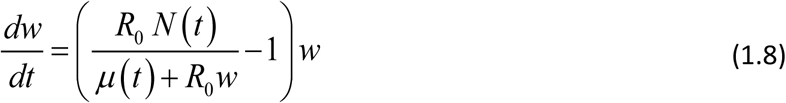

The key inputs for (1.7)-(1.8) is seasonal population density *N* (*t*), and stationary-mean *R*_0_ of rescaled system (Box 1). The role of conventional endemic equilibria (*w*^*^ = *y*^*^ = 1−1/ *R*_0_) is played by period solutions: {*w*^*^ (*t, a*)}, for (1.8), or {*w*^*^ (*t, a*), *y*^*^ (*t, a*)} for (1.7).

#### Comments

The above derivation of seasonal snail model (1.7) or (1.8) can be extended to any transmission system, e.g. MacDonald with mating (1.5)-(1.6), or SWB.

Besides seasonally varying snail population *N* (*t*), another source of variable transmission can be seasonal human-snail contact patterns, described by function *ω* (*t*). Indeed, both transmission coefficients *A, B* of the coupled system are proportional to *ω* (*t*). Two periodic inputs (*N* (*t*); *ω* (*t*)) are sometimes combined into a single product-type term (see e.g. ^17, 18^). While superficially ‘seasonal snail’ and ‘seasonal contact’ may look similar, there are significant differences in their equations, e.g. excess snail mortality in (1.7), (1.8); their dynamic responses may differ. In this paper we shall confine our analysis to seasonal snail population, and seasonal effect of periodic repeated MDA interventions.

### Dynamic snail population

For most parts of our analysis, we use prescribed seasonal snail function *N* (*t, a*), but molluscicide requires dynamic snail model, where populations {*N* (*t*), *y* (*t*)} could respond to any environmental or human-made change. Here we adopt a simplified version of snail model ^19^, where *N* (*t*) obeys a logistic growth model with prescribed maximal reproduction rate *β*, seasonal carrying capacity (CC) *K* (*t*), and mortality *ν*,

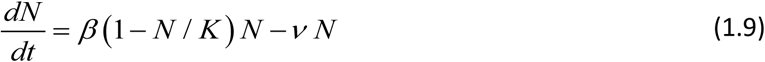

Periodic CC-function *K* (*t, a*) is taken as trigonometric (type I), peak (type II). Equation (1.9) has stable periodic solution *N* (*t, a*), which plays the role of the above prescribed type I-II *N*. We further assume snail infection has marginal effect on its growth/ mortality (due to small patency conversion fraction of snails). So equation (1.9) (variable *N*) is decoupled from transmission dynamics (1.7), (1.8), and can be used as input for MacDonald system.

Typical solution curves {*K* (*t, a*), *N* (*t, a*), *y* (*t*)}for type I-II seasonality are shown in Supplement (Figure ×). While function *K* (*t, a*) has its peak and trough at *t* = 0 and *t* = 1/ 2 (start, middle), *N* (*t, a*) lags behind (by about .1 of season), followed by *y* (*t*). We also note that seasonal mean values 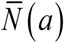 are now reduced 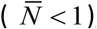, and their *R*_0_ (for comparison with prescribed N-case) must be adjusted accordingly.

## Results

### Seasonal variability and sustained infection

While stationary systems (1.2)- (1.3) allow direct analysis of equilibrium states via *R*_0_, non-stationary systems like (1.7)-(1.8) require numeric simulations. Here we examine the question of seasonal averaging, and single-host reduction for simple MacDonald system, via comparison of (1.2)-(1.7) vs. (1.3) -(1.8). Most results below involve periodic (endemic) equilibria *w*^*^ (*t, a*), and their seasonal mean 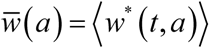. We list a few key results of our analysis (further details can be found in Supplement).

i. Stationary full (host-vector) and reduced (single host) MacDonald systems share the same equilibria and *R*_0_, but when perturbed from equilibrium (e.g. via MDA) their relaxation patterns to equilibrium differ, with reduced system relaxing faster (Supplement Figure A). Higher relaxation rate (rebound) would imply less efficient MDA control in reduced system compared to full MacDonald host-vector system.
ii. The role of stationary equilibria in variable environment is played by periodic (dynamic) solutions{*w*(*t, a*), *y* (*t, a*)}, see Figure 2 (more details shown in Supplement Figure C for trigonometric N, and Figure D for peak-type N.)
iii. Seasonal-mean values 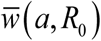 is a decreasing function of *a*, with maximum at the stationary endemic state (*a* = 0), where 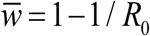. Depending on *R*_0_ the drop can become significant for high seasonality (*a* ≈ 1), up to 40% reduction. Reduced (single-host) model maintains similar qualitative patterns (loss of 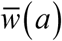 with *a*), but its sustained levels and seasonal means are higher, than the full MacDonald system (Figure 2(a)-Figure 3(a)).
iv. Seasonal variability, around its mean value, is relatively low for the worm burden *w*^*^ (*t*) (<5%), but attains much higher levels for infected snail density *y*^*^ (*t*). Peak worm burden *w*^*^ (*t*) lags behind peak snail density *N* (*t*) (and infected density *y* (*t*)) by about one quarter of a season.
v. Figure 2(b) and Figure 3(b) show similar features for type –II function *N* (*t*), in terms of its amplitude parameter *a*. Increased *a* (localized wet season) would lower periodic equilibria. As above, we observe marked difference between periodic patterns of reduced vs. complete MacDonald system (see Supplement Figure D). Their seasonal mean functions 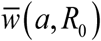 also depart significantly, particularly at large a, with complete 2D MacDonald predicting elimination at sufficiently high a (solid curves), while reduced model maintaining ‘positive’ endemic value for all a (dashed curves).
vi. We further extended this analysis by scanning the (*a, R*_0_) - parameter space to select regions of ‘endemicity’ 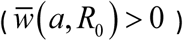, and ‘elimination’ 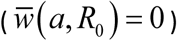. Thick red curve of Figure 4 marks the boundary between two regions, for trigonometric *N* (*t, a*) (left), and peak-type (right).

Overall, we find seasonal snail variability makes infection less sustainable (c.f. ^18^), and thus easier to control. The reduced host only system overestimates sustainability and control outcomes for MacDonald system.

**Figure 2:**
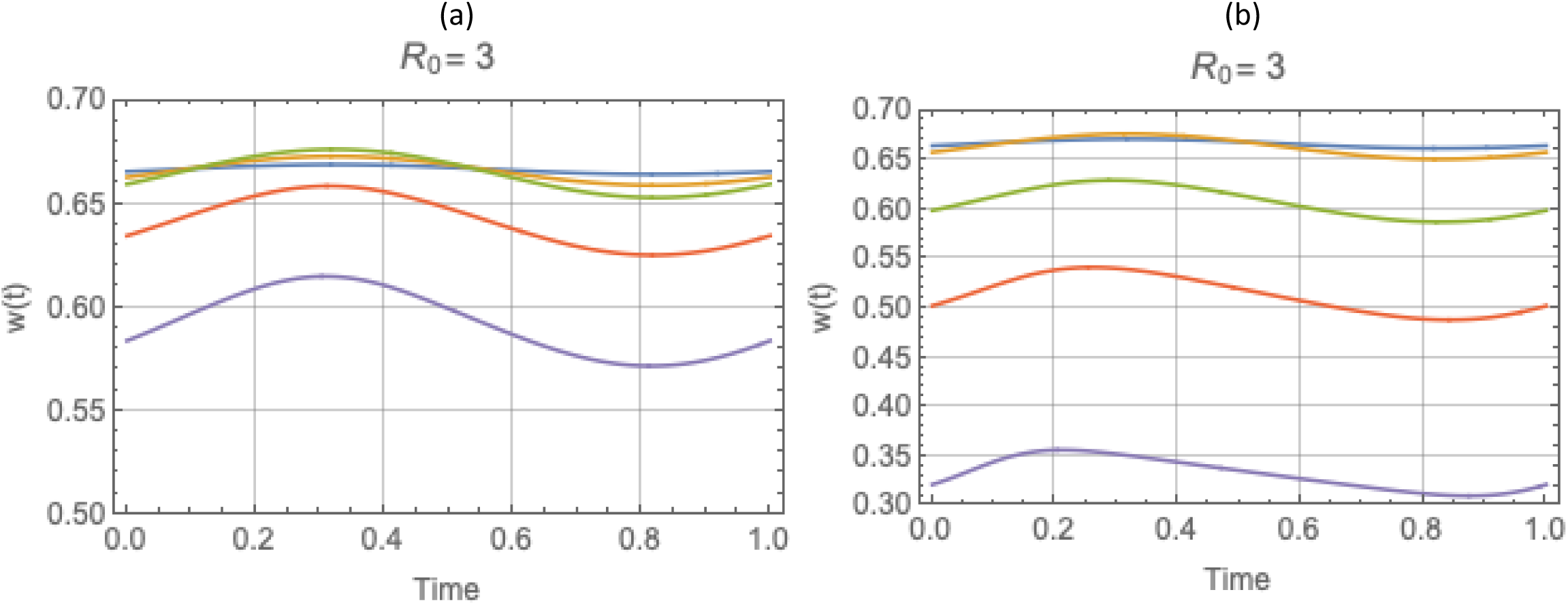
Periodic solutions *w*^*^ (*t*) of complete system: type I trigonometric ***N* (*t*)** (left panel) and type II peak ***N* (*t*)** (right panel), for values of ***R***_0_ = 3 and amplitude range 0 ≤ *a* ≤ 1, descending from stationary equilibrium *a* = 0 (blue) to ***a*** =.**95** (purple).

**Figure 3:**
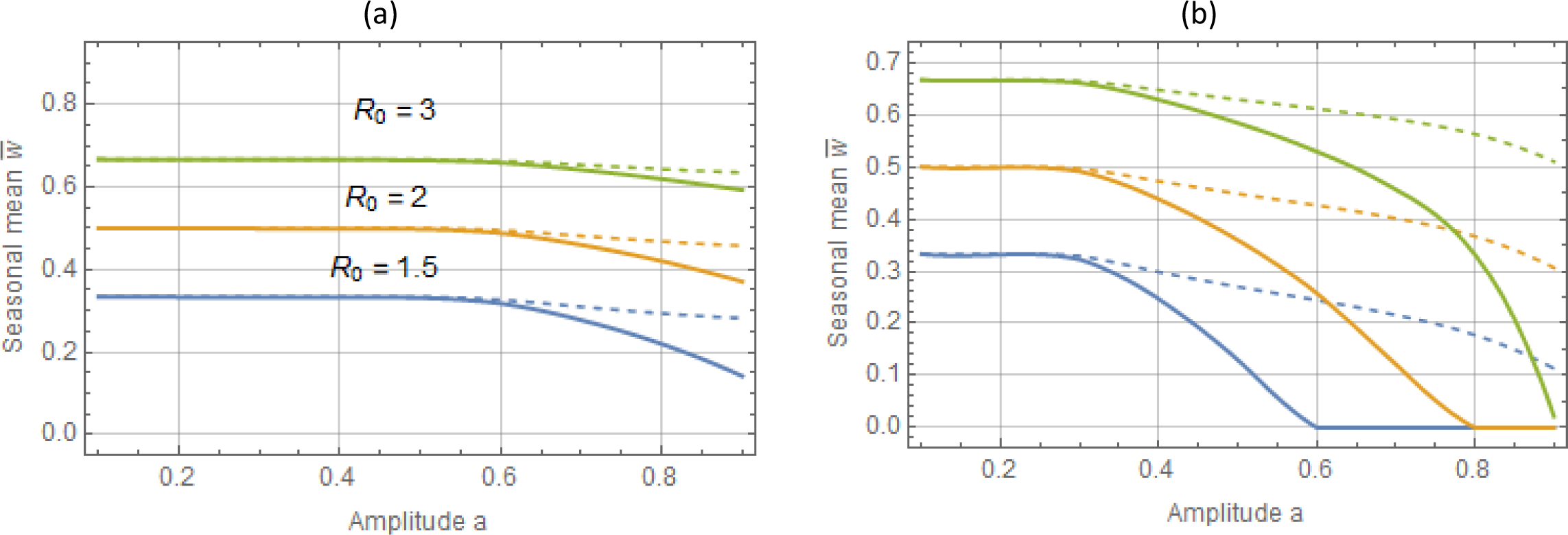
Seasonal mean 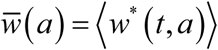 as functions of amplitude a : type I trigonometric N (t, a) panel (a), and type II peak – panel (b). The full human-snail MacDonald (solid) vs. reduced system (dashed) for 3 values R_0_ = 1.5; 2;3. Reduced model departs significantly from the full system at large seasonal amplitude (a > .6). It can thus over-predict seasonal mean burden by wide margin.

**Figure 4:**
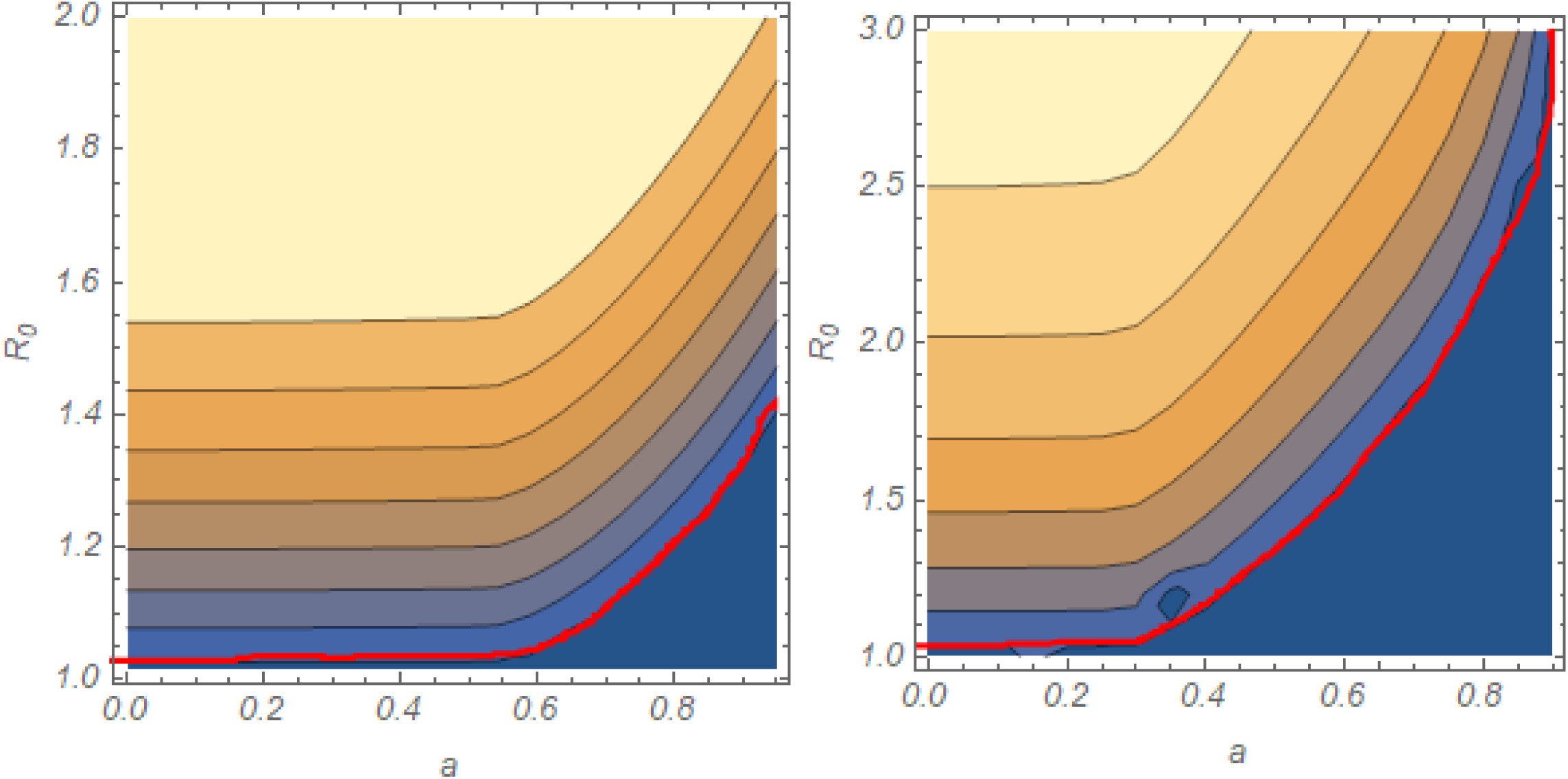
Iso-contours of seasonal mean MWB function 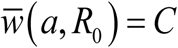 : left panel is trigonometric N (t, a), right panel – peak-type N (t, a). The range of contour values: 0 < C < 0.35 (left), and 0 < C < 0.6 (right). Thick red curve marks the boundary between endemic region (above), and zero-infection (below).

### MDA control in stationary environments

#### Periodic MDA in stationary environment for simple MacDonald system

An MDA session at time *T* is represented in our setup as instantaneous reduction of mean burden (due to short drug life-time), *w*(*T*) → *ε* _*f*_ *w*(*T*), where constant *ε f* combines drug efficacy *ε* (fraction of surviving worms), and population coverage fraction 0 < *f* < 1

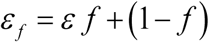

The latter applies to population groups (e.g. worm strata), or the entire community (MacDonald MWB). One can think of such MDA-event as a sharp spike of worm mortality over short duration. Formally represented by Dirac delta function.

Each MDA-event is followed by rebound - relaxation towards endemic state (equilibrium, periodic, et al). Such relaxation patterns can differ in complete (2D) vs. reduced MacDonald (Supplement Figure A). A regularly spaced sequence of MDA-events creates yet another type of periodic variability in dynamic system. This time periodicity is affected by worm mortality, rather than snail population growth/decay. So *γ* (*t*) becomes a periodic function with sharp (Dirac delta) spikes, or their finite approximation. As above, one can ask whether such periodic mortality *γ* (*t*) can be approximated by its ‘mean’ value,

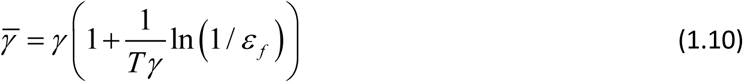

Here the natural *R*_0_ (Box 1) is replaced by effective reduced value due to (1.10)

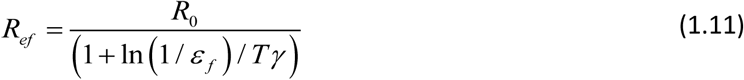

In particular, we ask whether reducing *R*_*ef*_ < 1 (via suitable combination of MDA frequency and efficacy-coverage) could lead to elimination. The condition for elimination in the stationary ‘mean’ system is

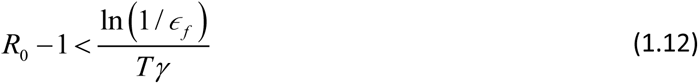

While numerator (1.12) is limited by drug efficacy (*ε f* ≤ *ε*) even at 100% coverage, its denominator could be made arbitrarily small by sufficiently frequent MDA sessions (small T).

To test the validity of MDA averaging, we run numeric simulations of two model, periodic MDA, and its ‘average model” with enhanced worm mortality (1.10). The results (Supplement Figure E) show elimination path for the mean-MDA system, predicted by *R*_*ef*_ < 1, while the exact MDA-response curve is locked in a periodic (limit) cycle pattern, even above critical frequency (*T* < *T*_*c*_). Such limit-cycle response patterns arise in many transmission models, including SWB (see ^23^), they can frustrate target reduction goals via repeated MDA regimen (see ^24^).

#### Parameter space analysis for MacDonald system with mating

Unlike simple MacDonald system (Box 1) with unstable-stable pair of equilibria (‘zero – endemic’), the mated system is bistable, for sufficiently high *R*_0_ > *R*_*c*_. Specifically, it has ‘zero’ and ‘endemic’ states (both stable), plus intermediate breakpoint (saddle) (Supplement Figure F). For detailed explanation, see (^1, 9^). Another important feature of mated MacDonald system, shared by SWB ^9^, is their dependence on two dimensionless parameters, e.g. 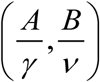, rather than single *R*_0_-their product.

The bistable nature (breakpoint) of mated MacDonald system has implications for MDA control. One such implication concerns regions in the (A, B) –parameter space which separate ‘endemic’ bistable state from ‘zero infection’. These regions deviate from isocontours of *R*_0_, so to predict elimination one needs two parameters (*A,B*), rather than single *R*_0_ (for details see Supplement Figure F, and ^9^).

Even more stark departures arise for dynamic MDA simulations. Here we can use stationary transmission environment, but vary transmission coefficients. The results (Supplement Figure G) show large diversity of outcomes, depending on transmission coefficients (A, B), proportional to population densities (*A* ∝ *N* - snail, *B* ∝ *H* - human). Different ‘snail per human’ ratios (A/B) correspond to different transmission environments (population densities), and under identical *R*_0_ they can produce different outcomes (Supplement Figure G). In some cases (lower ‘snail/human’ ratio), it goes to elimination after finite number of MDA cycles (though total duration could vary). In other cases (high ‘human/snail’ ratio), the system is locked in a limit cycle, due to post-MDA rebound.

We also observe a discrepancy between predictions of the reduced and complete MacDonald systems. Typically, reduced model would under-predict the MDA response, and show stronger rebound (Supplement Figure G).

The key conclusions are (i) conventional ***R***_0_ has little predictive value for endemic equilibrium analysis of mated MacDonald system (zero-to-endemic transition) ; (ii) reduced model (via snail quasi-equilibria) can grossly underestimate MDA responses; (iii) different choices of dimensionless (A,B) transmission coefficients that make up ***R***_0_ correspond to different snail-to-human abundance. High *A* (or ratio *N* / *H*) produce stronger post-MDA rebound, and compound time and effort required for elimination. Similar effect of snail environment on MDA outcomes was observed and studied for SWB transmission models (^23,25,26^). As with MacDonald systems, large snail-to-human ratio was shown to reduce the efficacy of repeated MDA. So near-identical host communities in terms of baseline infection, could produce vastly different MDA outcomes depending on snail environment (hot-spots vs. good responders).^26^

### Optimal MDA timing for seasonal transmission environment

In this analysis we used seasonal MacDonald system (1.7) with two types of seasonal *N* (*t*). In each case we computed its endemic periodic state {*w*^*^ (*t*), *y*^*^ (*t*)}, and run 6-year annual MDA with combined efficacy parameter *ε*_*M*_ = (1-*f*) + *ε f* = .4, corresponding to drug efficacy (*ε* = .15 − .25) and coverage fraction (*f* ≈ 0.75). For both types of seasonality *N* (*t,a*) we took *R*_0_ = 6; *a* = .95 (close to maximal amplitude).

The upper panels of Figure 5 compare MDA histories of seasonal *N* (*t*) vs. its stationary (seasonal-mean) counterpart. Both systems were treated at the season start (*τ* = 1, 2,×). In both cases, MDA outcomes (seasonal vs. mean stationary) appear close in terms of dynamic variable *w*(*t*), and its seasonal mean (Figure 5), over 6-year history.

**Figure 5:**
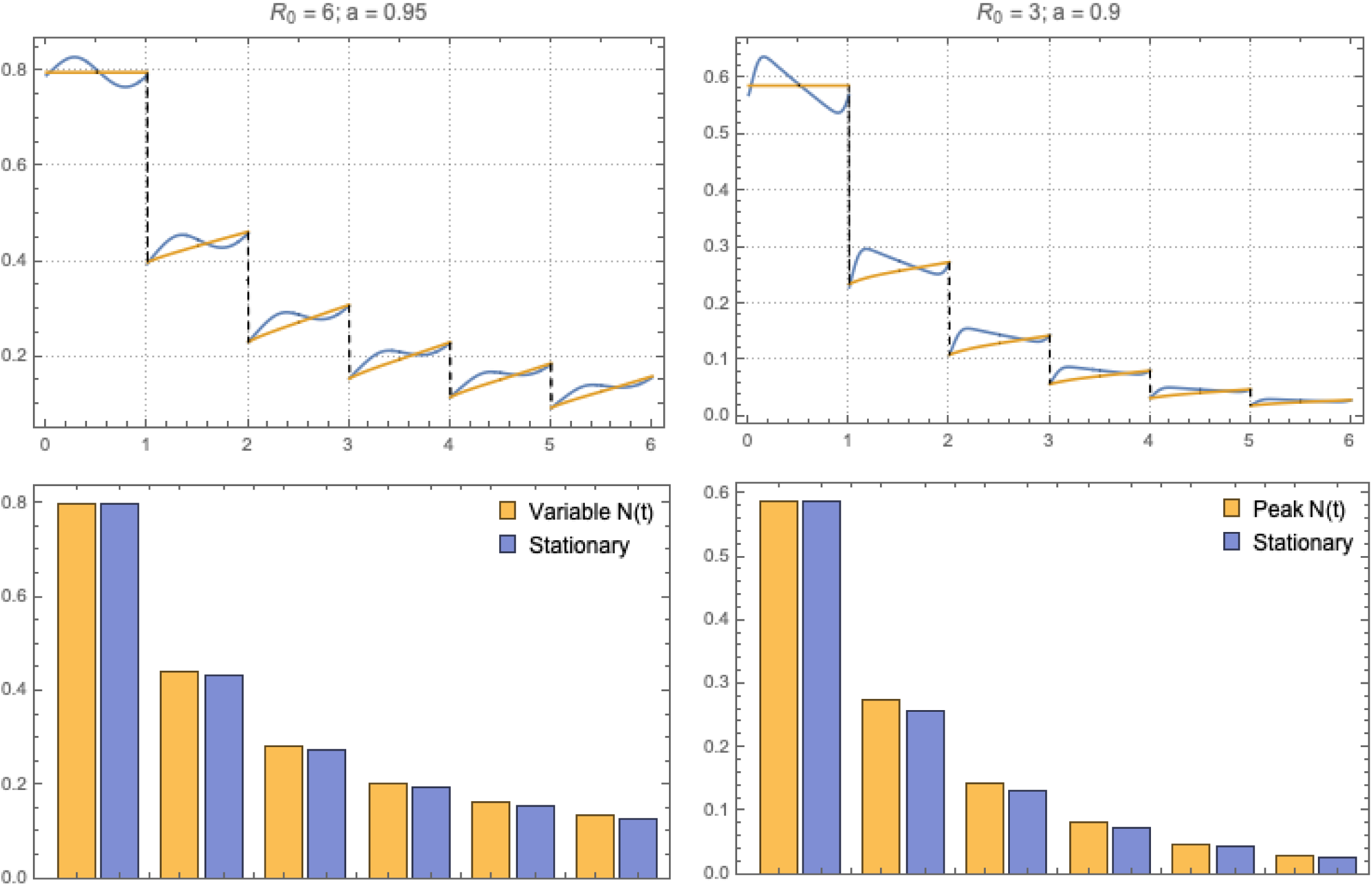
Annual MDA regimen for MacDonald system (1.7) with 2 types of seasonality and its adjusted stationary system: type I (left column) and type II (right column). Top plots compare MDA histories of seasonal MacDonald (blue) vs. the corresponding stationary (seasonal mean) MacDonald (yellow). Bottom panels show their seasonal-mean burden on tears 1-6.

However, different seasonal timing *τ* of MDA can produce marked differences between stationary and non-stationary models. Upper panels of Figure 6 demonstrate this effect by comparing MDA histories of *τ* = 0 (season start) vs. *τ* = .5 (middle). It raises the question of *optimal choice* of MDA timing, to achieve maximal burden reduction by the end of the program (Y6).

**Figure 6:**
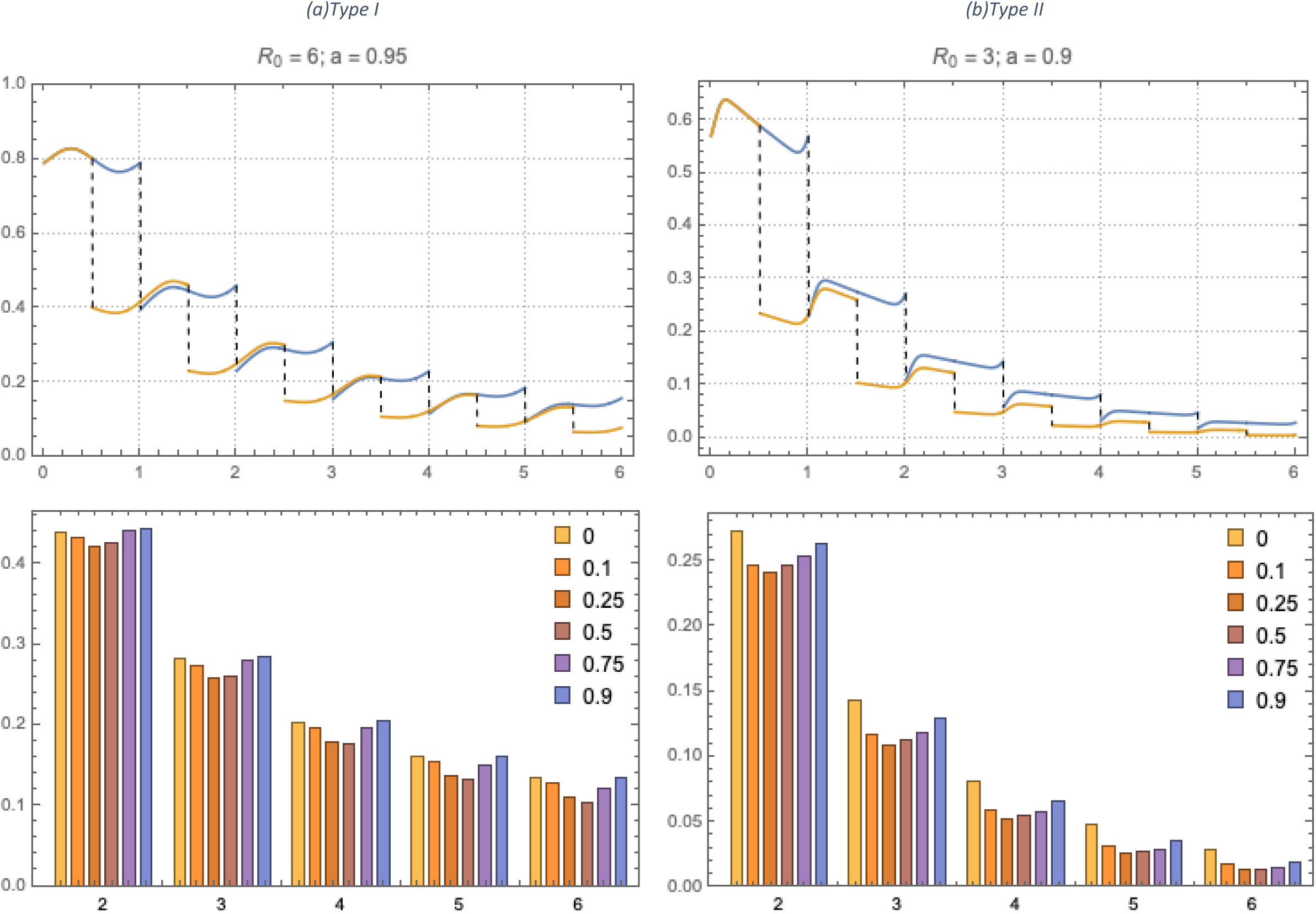
Seasonal timing (0 ≤ *t*_*S*_ < 1) of annual MDA for both types of seasonal MacDonald systems. Upper panel show 2 dynamic histories at *t*_0_ = 0 (blue) and shifted *t*_0_ = .5 (yellow). The low panels seasonal mean of each timing regimen *t*_*S*_ (average *w*(*t*) over post treatment interval (*t*_*S*_, *t*_*S*_ +1)).

To this end, we run multiple simulations with different choices (0 < *τ* < 1), relative to snail peak (*τ* = 0). In each case, we estimated mean effect of a given *τ* - strategy, by averaging over the proper time interval [*τ, τ* +1].

Figure 6 shows the results (mean w – values) over 6-year control history for different choices of time shift *τ*. We find overall effect of *τ* could be significant between ‘best’ and ‘worst’ timing, up to 40% improvement (Y6 over Y1) for trigonometric N and to 50% - for peak N. In both cases, we took high amplitude seasonality (*a* = 0.95). We observe the optimal timing *τ* is between one quarter and half-season for type I, and close to a quarter for type II. Here season start is identified with peak snail population. The results are summarized in Table 2, for each *τ* we compute 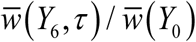 - endemic.

**Table 1:** Relative drop of seasonal mean burden as function of amplitude 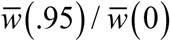, for 3 BRN values R_0_ = (1.5, 2.5, 4).

|  | 1.5 | 2.5 | 4 |
| --- | --- | --- | --- |
| Reduced | 0.84 | 0.93 | 0.96 |
| Complete 2D | 0.27 | 0.81 | 0.92 |

**Table 2:** Progress (Y6/Y1) for several different choices of MDA timing τ of Figure 6. Optimal timing is close to τ = 0.5 past snail peak.

| $\tau$ | 0 | 0.1 | 0.25 | 0.5 | 0.75 | 0.9 |
| --- | --- | --- | --- | --- | --- | --- |
| I | 0.304 | 0.295 | 0.261 | 0.24 | 0.275 | 0.3 |
| II | 0.103 | 0.0687 | 0.0519 | 0.0535 | 0.0551 | 0.0702 |

### Optimal timing for molluscicide and integrated strategies

Molluscicide is considered a viable option for control of schistosomiasis ^27^. In this section we explore optimal timing for snail control (molluscicide), and integrated strategy (MDA + molluscicide), using logistic snail model (1.9) coupled to MacDonald system (1.7).

Snail control (molluscicide) is implemented similar to MDA, as instantaneous event, whereby dynamic variables {*N* (*t*), *y* (*t*)} are reduced by snail survival fraction (0 < *ε*_*S*_ < 1) – the efficacy of molluscicide, and implementation time *τ*,

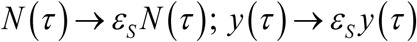

We run control simulations for both types of seasonality with large amplitude factor, *a* = .9 (type I), and *a* = .6 (type II). In all cases, snail mortality and maximal growth were fixed at *ν* = 4 /year, *β* = 20 /year, consistent with estimated parameters ^19^. We also fixed *R*_0_ = 6, though its effective values (vs. fixed-N case) are somewhat lower, reduced by factor 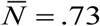 (type I), and 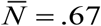 (type II).

As above we sample several choices of seasonal timing (0 ≤ *τ* < 1), and for each *τ* -strategy compute its seasonal mean value.

Figure 7 shows typical 6-year histories for integrated control (MDA + molluscicide) for both types of seasonality, CC-function *K* (*t, a*). Two timing choices are compared, suboptimal *τ* = 0 (blue), and *τ* = 0.5 (yellow), close to optimal. In both cases, we took high seasonal variability (*a* = 0.9 for type I, and *a* = 0.6 for type II). Application of repeated molluscicide rapidly brings snail system to a periodic pattern (after 1-2 rounds) for function *N* (*t*) - total population (unaffected by MDA). But infected snail density *y* (*t*) approaches zero (due to MDA effect on snail infection).

**Figure 7:**
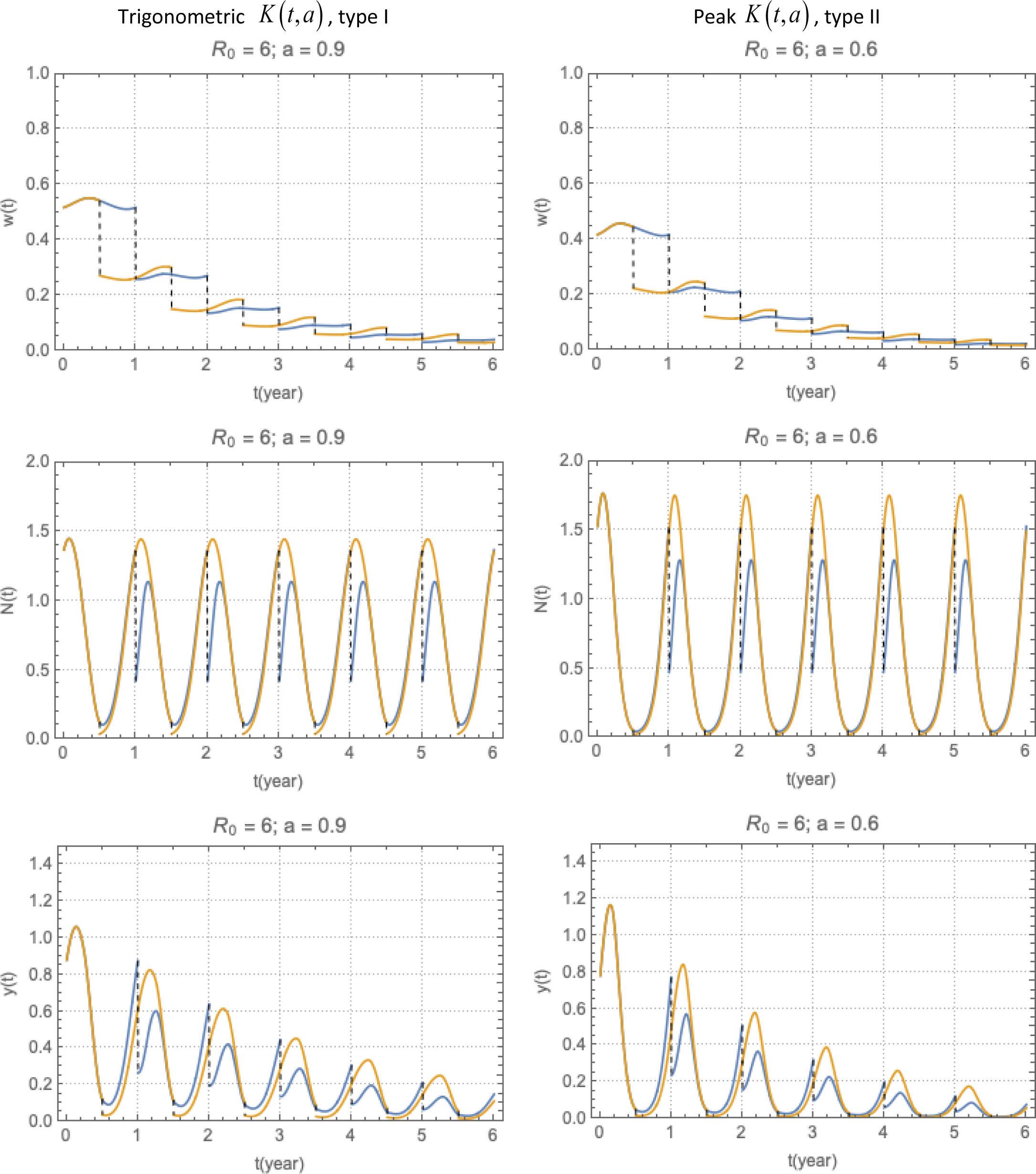
Combined MDA + molluscicide strategies for type I-II seasonality (left-right panels). Two curves correspond to start of season (suboptimal timing τ = 0), and mid-season (τ = 0.5, close to optimal).

Next we discuss specifically optimal timing to two types of seasonality. For *molluscicide* alone the results are shown in Table 3 (see supplement). The optimal timing (maximal MWB reduction) falls near start of season (*τ* ≈ 0) where CC-function *K* (*t, a*) reaches its maximum. The overall progress (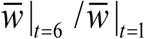 varies in the range 8-30% reduction of MWB, depending on the type of seasonality, and clearing efficiency *ε* _*S*_ (surviving snail fraction).

**Table 3:** Progress of molluscicide control (Y6/Y1) at several values of seasonal timing 0 ≤ τ < 1, and molluscicide efficacy ε_S_ = {0.5, 0.3, 0.1}.

| Type I |  | 0 | 0.1 | 0.25 | 0.5 | 0.75 | 0.9 |
| --- | --- | --- | --- | --- | --- | --- | --- |
| Amplitude | 0.5 | 0.915 | 0.922 | 0.959 | 0.985 | 0.945 | 0.923 |
| $a = 0.9$ | 0.3 | 0.863 | 0.877 | 0.935 | 0.973 | 0.905 | 0.872 |
|  | 0.1 | 0.771 | 0.804 | 0.9 | 0.936 | 0.817 | 0.774 |
| Type II |  | 0 | 0.1 | 0.25 | 0.5 | 0.75 | 0.9 |
| Amplitude | 0.5 | 0.885 | 0.901 | 0.967 | 0.984 | 0.927 | 0.897 |
| $a = 0.6$ | 0.3 | 0.818 | 0.846 | 0.946 | 0.971 | 0.873 | 0.83 |
|  | 0.1 | 0.705 | 0.762 | 0.916 | 0.926 | 0.754 | 0.705 |

The *MDA-only* strategy looks qualitatively similar to the previous case, prescribed N-function (Figure 6). The optimal timing is close to mid-season *τ* ≈ .5 (Table 4 and supplement) is near mid-season. The mean burden reduction is much more significant now, varies in the range 80-97%, compared to molluscicide.

**Table 4:** Progress (Y6/Y1) for MDA alone for several values of seasonal timing 0 ≤ τ < 1, and 3 levels of MDA efficacy ε_M_ = {0.3, 0.45, 0.5}}.

| Type I |  | 0 | 0.1 | 0.25 | 0.5 | 0.75 | 0.9 |
| --- | --- | --- | --- | --- | --- | --- | --- |
| Amplitude | 0.3 | 0.0921 | 0.102 | 0.09 | 0.0537 | 0.0557 | 0.0751 |
| $a = 0.9$ | 0.45 | 0.201 | 0.208 | 0.191 | 0.154 | 0.161 | 0.185 |
|  | 0.5 | 0.246 | 0.251 | 0.234 | 0.199 | 0.209 | 0.231 |
| Type II |  | 0 | 0.1 | 0.25 | 0.5 | 0.75 | 0.9 |
| Amplitude | 0.3 | 0.0571 | 0.0687 | 0.0495 | 0.0297 | 0.0308 | 0.0416 |
| $a = 0.6$ | 0.45 | 0.152 | 0.163 | 0.136 | 0.109 | 0.114 | 0.132 |
|  | 0.5 | 0.195 | 0.206 | 0.177 | 0.151 | 0.157 | 0.176 |

The *combined* (MDA+ molluscicide) strategy was implemented at fixed snail removal rate, *ε*_*S*_ = 0.3, and 3 choice of MDA efficacy-coverage, *ε*_*M*_ = (0.3, 0.45, 0.5). In the first experiment we run simultaneous MDA and molluscicide with different seasonal timing *τ*. We also looked at then effect of seasonal amplitude *a*, strong or weak seasonality. The results are shown in Table 5. Overall, the effect of seasonal timing *τ* is less significant now, compared to molluscicide or MDA alone, though it increases with higher MDA efficacy. We believe this is due to different optimal values of *τ* for MDA (typical *τ* ≈ 0.5), and for molluscicide (typical *τ* ≈ 0).

**Table 5:** Combined MDA + molluscicide strategy conducted simultaneously (timing τ = {0; 0.1; 0.25;×}) for fixed molluscicide efficacy ε_S_ = 0.3, and 3 choices of MDA efficacy ε_M_ = (0.3, 0.45,.05).

| Strong seasonality | Type I<br>Amplitude<br>$a = 0.9$ | | 0 | 0.1 | 0.25 | 0.5 | 0.75 | 0.9 |
| --- | --- | --- | --- | --- | --- | --- | --- | --- |
|  |  | 0.3 | 0.0299 | 0.0324 | 0.0396 | 0.0358 | 0.0295 | 0.0289 |
|  |  | 0.45 | 0.0998 | 0.103 | 0.121 | 0.125 | 0.108 | 0.101 |
|  |  | 0.5 | 0.135 | 0.139 | 0.16 | 0.169 | 0.148 | 0.138 |
| | Type II<br>Amplitude<br>$a = 0.6$ | | 0 | 0.1 | 0.25 | 0.5 | 0.75 | 0.9 |
|  |  | 0.3 | 0.0188 | 0.0223 | 0.028 | 0.0233 | 0.0178 | 0.0173 |
|  |  | 0.45 | 0.0714 | 0.0785 | 0.0974 | 0.095 | 0.0759 | 0.0707 |
|  |  | 0.5 | 0.1 | 0.109 | 0.134 | 0.134 | 0.109 | 0.101 |
| Weak seasonality | Type I<br>Amplitude<br>$a = 0.5$ | | 0 | 0.1 | 0.25 | 0.5 | 0.75 | 0.9 |
|  |  | 0.3 | 0.0531 | 0.0544 | 0.0572 | 0.0584 | 0.0536 | 0.0526 |
|  |  | 0.45 | 0.146 | 0.147 | 0.152 | 0.16 | 0.153 | 0.148 |
|  |  | 0.5 | 0.187 | 0.187 | 0.193 | 0.205 | 0.196 | 0.19 |
| | Type II<br>Amplitude<br>$a = 0.3$ | | 0 | 0.1 | 0.25 | 0.5 | 0.75 | 0.9 |
|  |  | 0.3 | 0.0511 | 0.053 | 0.0563 | 0.0573 | 0.0516 | 0.0504 |
|  |  | 0.45 | 0.142 | 0.144 | 0.15 | 0.159 | 0.15 | 0.144 |
|  |  | 0.5 | 0.183 | 0.184 | 0.191 | 0.204 | 0.194 | 0.186 |

The maximal reduction varies (depending on *ε*_*M*_) in the range 90-97% for type I (*a* = 0.9), and somewhat better results for type II with (*a* = 0.6); both cases represent strong seasonal variability. We observe similar results, but significant reduction (82-95%) for moderate or weak seasonality (*a* = 0.5, type I, and *a* = 0.3 type II). The reason for improved control outcomes at higher *a*, is the reduced sustainability (endemicity) in highly variable environments, as explained above.

In the final experiment we took independent optimal *τ*, suggested by earlier simulations, *τ* = 0.5 for MDA, and *τ* = 0 for molluscicide. The results are shown in Table 6 (upper half), and compared results to the previous case, i.e. simultaneous MDA+ molluscicide schedule Table 5.

**Table 6:** Optimal MDA+molluscicide control for two types of seasonality at different seasonal amplitude a and 3 choices of MDA efficacy ε_M_ = (0.3, 0.45,.05). Optimal MDA was run at the midseason (τ = 0.5), while optimal molluscicide at the start of season (τ = 0).

|  |  | Type I |  | Type II |  |
| --- | --- | --- | --- | --- | --- |
| | | $a = 0.9$ | $a = 0.5$ | $a = 0.6$ | $a = 0.3$ |
| Independent | 0.3 | 0.010 | 0.030 | 0.005 | 0.028 |
|  | 0.45 | 0.048 | 0.090 | 0.030 | 0.088 |
|  | 0.5 | 0.072 | 0.123 | 0.049 | 0.118 |
| Synchronous | 0.3 | 0.029 | 0.053 | 0.017 | 0.050 |
|  | 0.45 | 0.100 | 0.146 | 0.071 | 0.142 |
|  | 0.5 | 0.135 | 0.187 | 0.100 | 0.183 |

Overall, we achieve further improvement (by factor 2-3) compared to best selection of Table 5 (lower half of Table 6). Depending of seasonal amplitude/type, and MDA coverage-efficacy *ε*_*M*_, the reduction can go as low as 0.5% of the baseline MWB.

We also find that adding molluscicide to MDA can bring a significant improvement, when both controls are properly timed.

## Conclusions and discussion

The Basic reproduction number *R*_0_ is a dimensionless parameter widely used in infection modeling. It applies to model reduction (via non-dimensionalization), analysis of equilibria, endemic states and control. In simple cases it can be estimated directly from model inputs (transmission, recovery/ loss rates), and provides full information on endemicity or elimination. One can extend it for large systems with multiple compartments, like SWB, but its scope is more limited. In the SWB models, analysis of endemicity/ elimination requires at least 2 dimensionless parameters (see ^9^).

Variable dynamic environment poses significant challenges. Here ***R***_0_ alone cannot be used for prediction / analysis, without additional information on temporal variability. A conventional approach of seasonal averaging and reduction to quasi-stationary model is approximately valid at low seasonal variability. But it can depart markedly at high seasonal amplitude, both in terms of sustained infection and MDA responses. In general, increased variability makes infection less sustainable, so seasonal-mean MWB would drop with increased amplitude (or standard deviation) of varying snail population. It implies that such system may be more amenable to control interventions, compared to its stationary (mean) counterpart.

Regular MDA regimen gives another example of periodic dynamic perturbation, where naïve application of *R*_0_ could also fail. Comparing two types of variability, *seasonal snail* vs. *repeated MDA*, we draw the following conclusions: i) in the former case (seasonal snail), *R*_0_ would typically overestimate sustainability of infection; ii) in the latter case, if we assume suitable drug efficacy and MDA coverage, control-induced reproductive number *R*_*c*_ would predict elimination, while the system is locked in a limit cycle.

One basic conclusion of our analysis is the need to employ direct dynamic simulations in all cases of unsteady (seasonal) transmission.

Such direct simulation allows one to explore, among other optimal timing for MDA, snail control or integrated strategies in variable environment.

For MDA alone we found such optimal timing is close to half-season, which corresponds to minimal carrying capacity for snails. It’s also close to minimal snail density, which could be used as more practical measure for optimal MDA timing. We also found its progress (relative burden reduction), over short-term 6-year control program, could vary markedly, and ‘optimal’ timing can achieve up to 50% improvement compared to suboptimal’ timing.

For molluscicide control, the optimal timing is shifted closer to start of season, i.e. snail peak, but overall effect of molluscicide on burden reduction is less significant than MDA, even at high killing efficacy.

The combined strategy MDA and molluscicide, can give an additional reduction of worm burden, compared to MDA alone. When both interventions are implemented simultaneously, intra-seasonal variability is less significant, since optimal choices for each one differs. But each intervention implemented at its own optimal timing can achieve much better results.

Overall, snail control can add significantly to the program outcome; in some cases it can lead to near-elimination after short (6-year) duration

High seasonal variability gives better control outcomes in all cases, as it makes transmission and endemicity less sustainable.

## Data Availability

No data used.

## Supplement

### SWB system

The SWB approach was developed and refined in several papers (^8-10,22-25^). It has many advantages compared to MacDonald MWB, simple or ‘mated’ (NB) version. SWB system carries detailed information on burden distribution and egg release by host population, but it requires no prior assumptions on burden distribution, like NB. Within-host worm biology (mating, mortality, fecundity) is naturally accommodated in SWB. While the number of SWB variables can be large (depending on stratification), it has the same parameters as MacDonald. In fact, the MWB variable *w*(*t*) is given by the first statistical moment (mean) of SWB distribution, and the *w*-equation of (1.7)-(1.4) is derived from SWB equations. The main difference between MWB and SWB comes in the snail force of infection (FOI) Λ -. For MacDonald system, Λ is a function of single (MWB) variable *w*, for SWB it depends on human infectivity *E* - combined egg release by worm burden strata, worm mating and fecundity. There is no direct link between *w* and *E*.

Like simple MacDonald system, coupled SWB-snail system has a *R*_0_ parameter that can be computed from its basic inputs. Such a parameter has similar meaning to MacDonald *R*_0_, namely 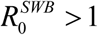 1implies unstable zero and stable endemic state, while 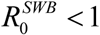 has stable zero. Beyond that it carries no information on endemic state, variable environment, or dynamic interventions (MDA control outcomes).

In fact, more detailed information on SWB equilibria can be derived from two transmission confidents *A, B* rather than their product (like *R*_0_). So, communities with identical human infection, but different snail environment (hence different pairs *A, B*) can produce vastly divergent control outcomes (Supplement).

### Parameter *R*_0_ of coupled SWB-snail system

Here we derive *R*_0_ formula for coupled SWB-snail system, assuming logistic snail population growth, and SEI snail infection dynamics, and nonlinear snail force of infection (FOI) (^29^).

The relevant input parameters are listed in Table 1.

**Table 1:** input parameters for coupled SWB-snail model

|  |  |
| --- | --- |
| $\Delta w > 1$ | SWB worm step (threshold for mating) |
| $\rho_0$ | max egg-release/mated female worm |
| $\rho_m = \rho_0 z^m$ | Female worm fecundity in m-th worm burden stratum with crowding factor ( $z < 1$ ) |
| $\phi_m = \frac{m}{2} \left[ 1 - 2^{-m} \binom{m}{m/2} \right]$ | Mean mated couple count for m-stratum (m adult worms) |
| $\gamma, \mu$ | Worm mortality and host population turnover |
| $a$ | “probability of worm establishment/contact / snail” |
| $\omega$ | Human-snail contact rate |
| $b$ | probability of snail invasion by merozoite |
| $N$ | snail density/unit habitat |
| $H$ | human population density/unit habitat |
| $c$ | snail patency conversion fraction |
| $\nu$ | snail mortality |
| $r$ | patency conversion rate |
| $\beta$ | Maximal growth rate (for logistic snail population) |
| $A = a \omega N$<br>$B = b \omega H \rho_0$ | Transmission coefficients: snail-to-human and human-to-snail |
| $\frac{c(1-\nu/\beta)}{(1+\nu/r)}$ | snail contribution to BRN of the coupled system |

The complete *R*_0_ consists of 3 factors: (i) conventional Ross-MacDonald *R*_0_, expressed through transmission coefficients A,B, and worm, snail, host mortality/turnover rates; (ii) SWB factor, Formula applies to coupled SWB-snail system with simple SI snail, like. For modified SEI snail used in the current version 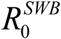 is augmented by an additional “snail factor”:

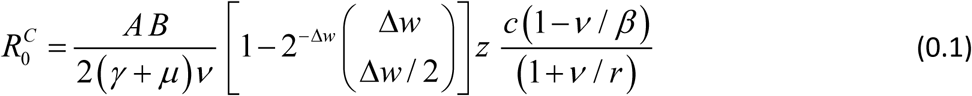

Formula (0.1) could be further extended to demographically structured SWB communities (e.g. children + SAC + adult et al).

Formula (0.1) can be derived from the Jacobian analysis of the coupled system at zero equilibrium. Specifically, condition *R*_0_ = 1 marks a transition from stable to unstable zero (i.e. stable endemic state). But there is no direct link between *R*_0_ and endemic state, or possible MDA control outcomes.

In fact, a wide range of equilibria arises by varying dimensionless transmission rates {*A* / (*γ* + *μ*); *B* /*ν* } (e.g. different human-to-snail population density *H* / *N*), with different MDA responses.

### Modeling setup

Relaxation patterns to endemic equilibria for reduced and complete MacDonald system

**Figure A:**
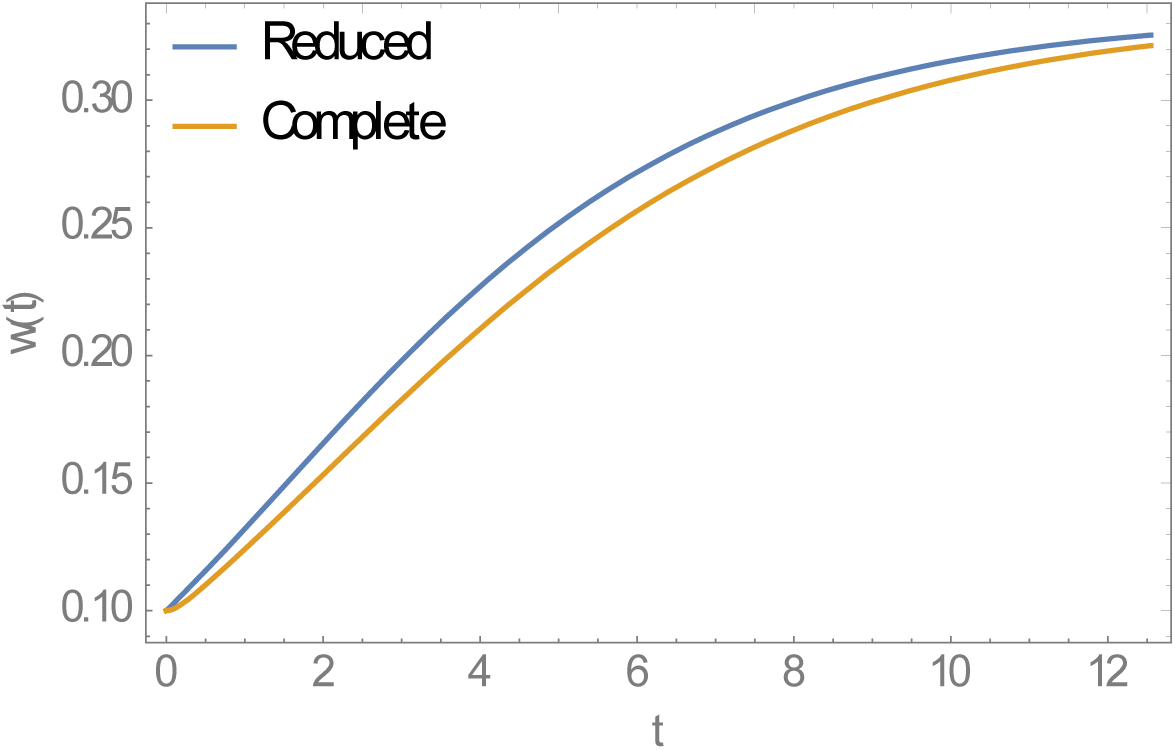
Relaxation patterns of stationary MacDonald systems (reduced model, full model). Both systems relax to the same equilibrium w^*^ = 1−1/ R_0_, but reduced model predicts faster relaxation rate.

Typical dynamic seasonal snail patterns for MacDonald system with CC *K* (*t, a*), with properly adjusted snail mortality, so that *y* (*t*) < *N* (*t, a*), for all t.

**Figure B:**
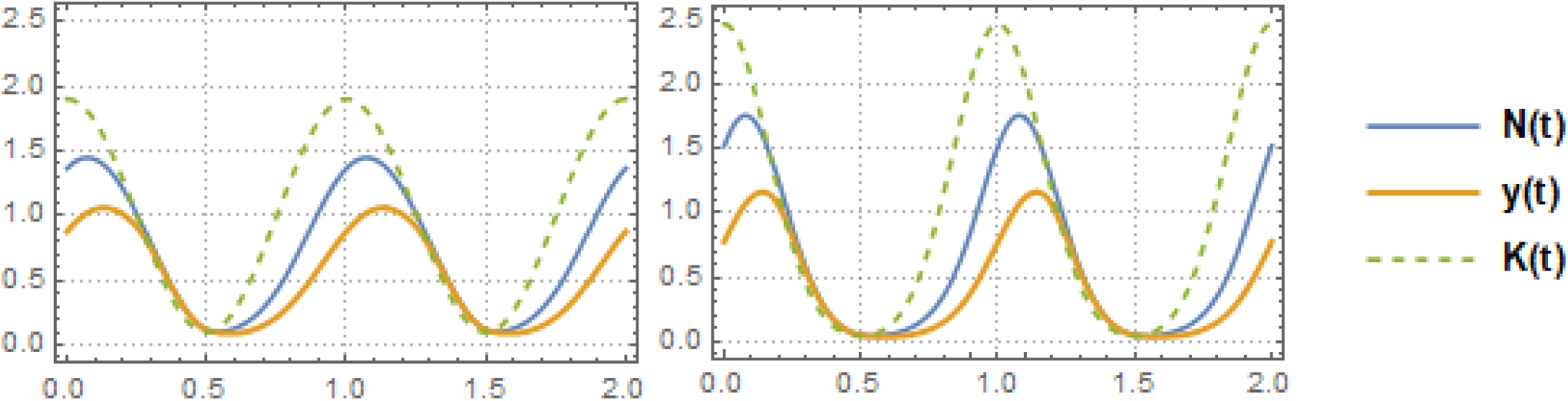
Typical seasonal patterns for logistic snail model with CC-function K (t, a), relative growth rate β /ν = 5, and snail mortality ν = 4 /year. Left panel show shows MacDonald solution {N (t), y (t)} for trigonometric K (t, a) of amplitude a = 0.9, R_0_ = 6. Right panel does the same for peak-type K (t, a) with a = 0.6; R_0_ = 6. Seasonal mean values 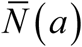 are reduced compared to prescribed N-case: 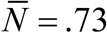 (type I, left panel), and 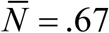 (type II, right panel)

### Seasonal variability

**Figure C:**
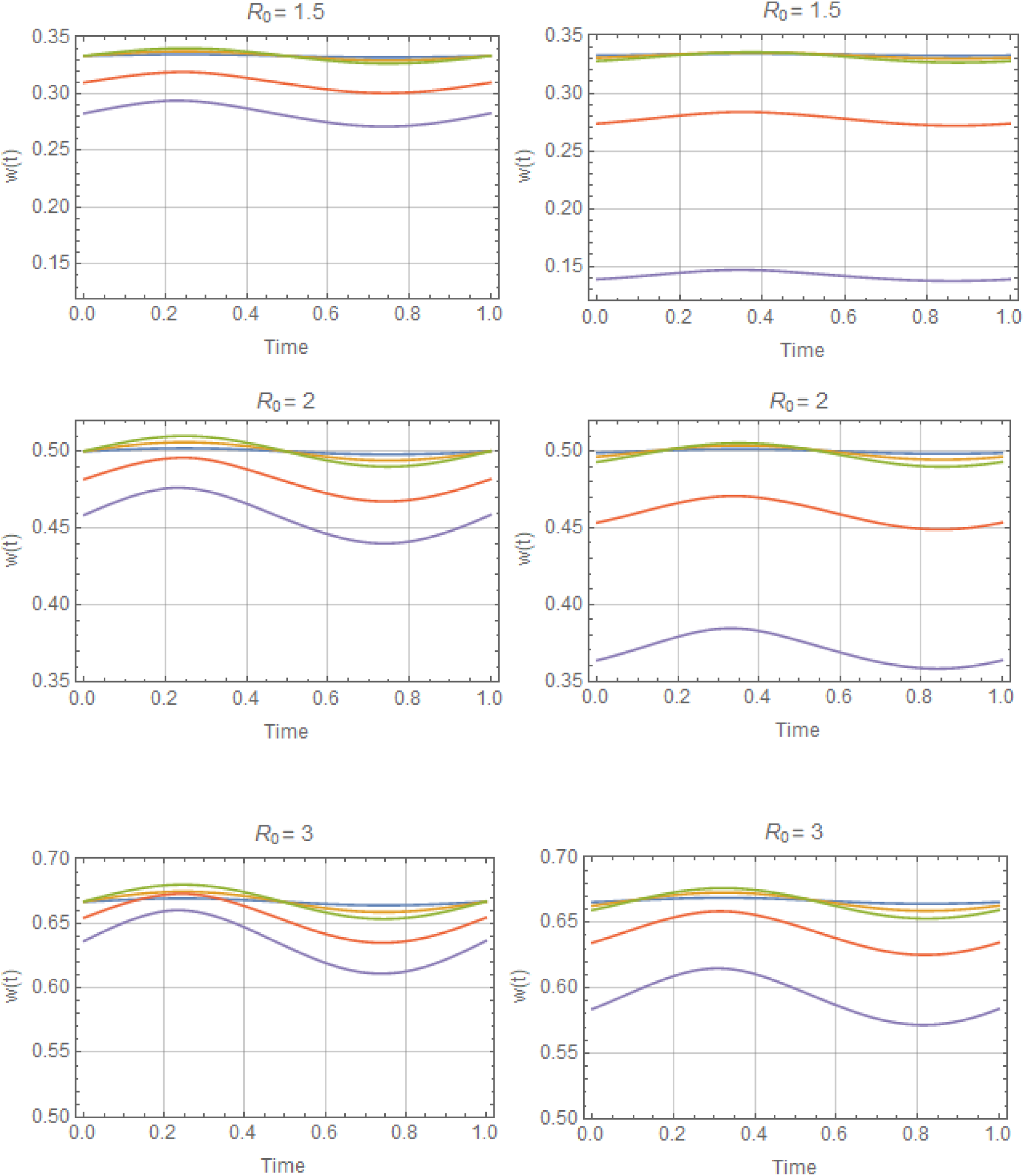
Periodic solutions *w*^*^ (*t*) with trigonometric *N* (*t*), for values of *R*_0_ = (1.5, 2, 3) and amplitude range 0 ≤ *a* ≤ 1, descending from stationary equilibrium *a* = 0 (blue) to *a* = .95 (purple). Reduced system (left), and complete system (right). Variation of *w*^*^ (*t*) about its mean 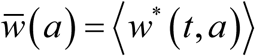 is about 5%-10%, for all (*R*_0_, *a*). But seasonal mean function 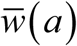 drops below its stationary equilibrium value *w*^*^ = 1−1/ *R*_0_, for all *a* > 0.

**Figure D:**
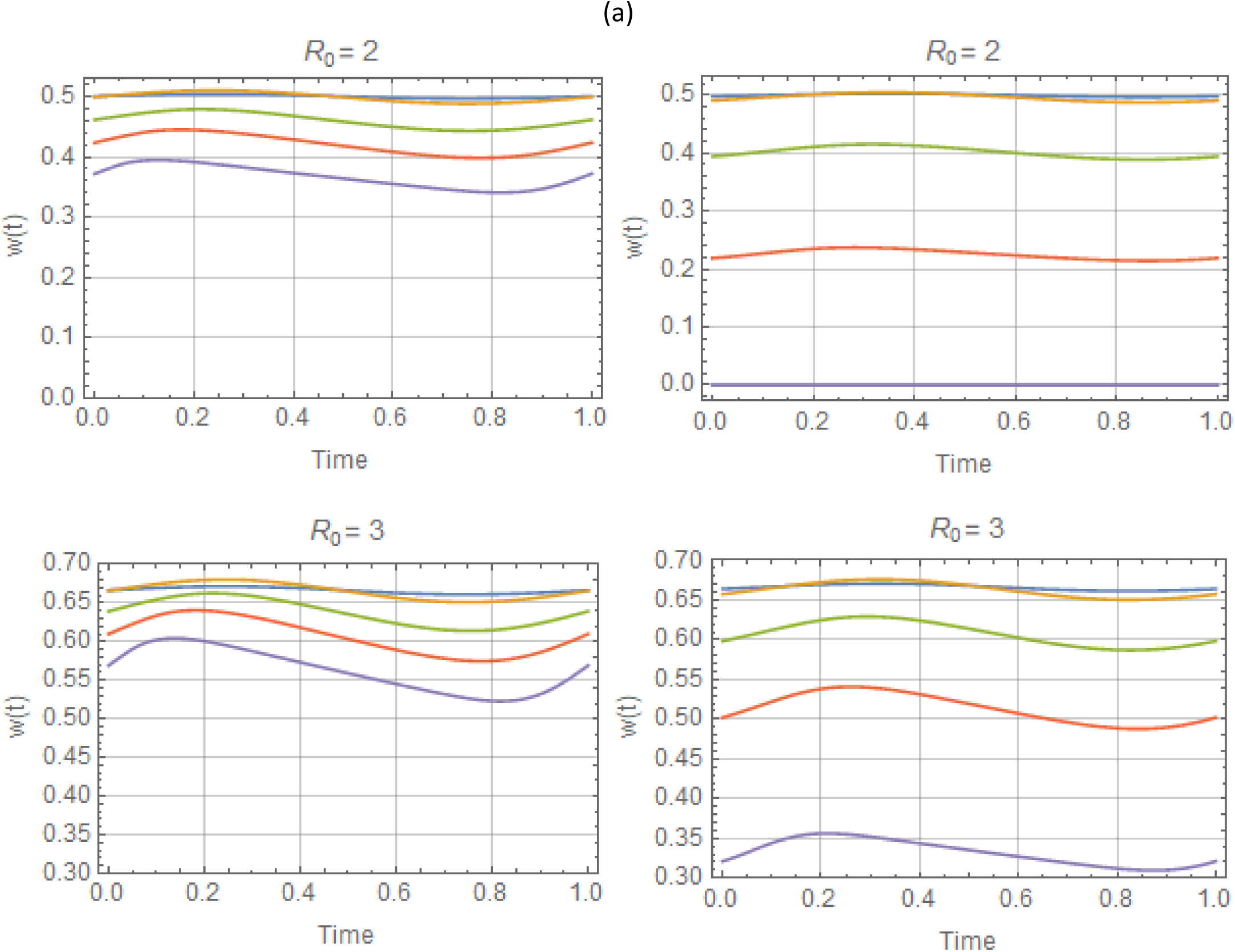
Periodic solutions *w*^*^ (*t, a*) for –peak-type *N* (*t* | *a*), with *R*_0_ = 2, 3, and a range of amplitude values 0 ≤ *a* ≤ .8 (top to bottom). As in Figure C, left column shows reduced system, right column - complete system.

### MDA control in stationary environments

Comparison of periodic MDA and its average-model prediction for simple MacDonald The results (Figure E) show elimination path (yellow curve) for the mean-MDA system, predicted by *R*_*ef*_ < 1, while the exact MDA-response curve (blue) was locked in a periodic (limit) cycle pattern, even above critical frequency (*T* < *T*_*c*_).

**Figure E:**
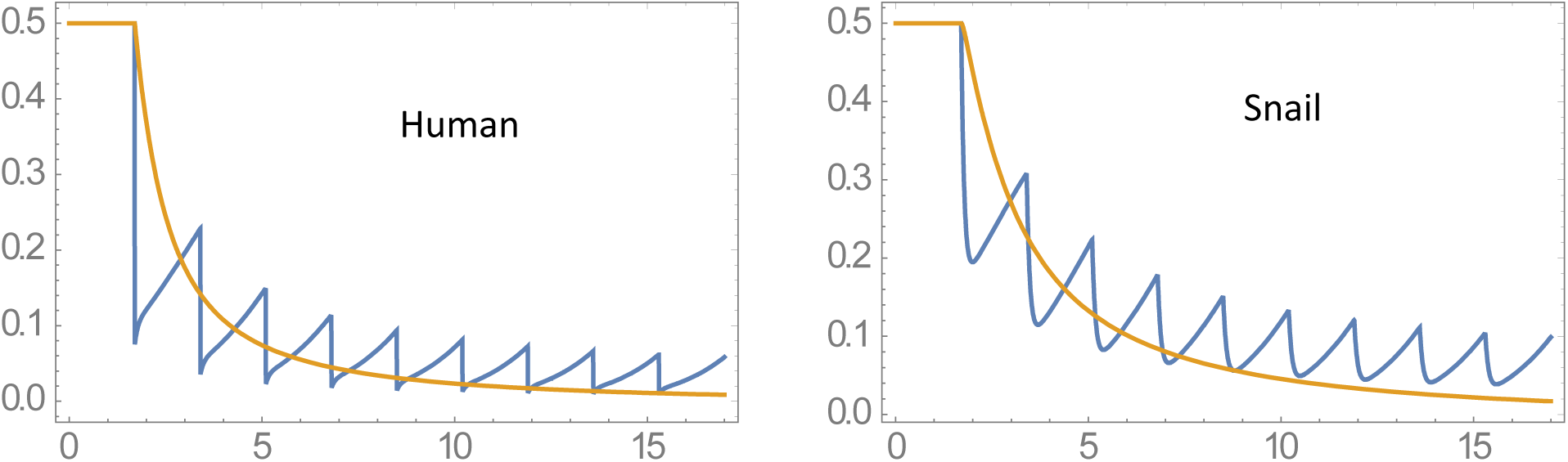
Averaging effect on MDA: two panels compare repeated periodic MDA history (blue), and the corresponding ‘mean drug clearing’ model (yellow). The former approaches limit cycle regime, the latter goes to elimination (effective BRN R_ef_ < 1). Here we have chosen treatment period T = 1.7 < T_c_ = 1.89.

### Parameter space analysis for mated NB MacDonald

MacDonald system with mating and NB worm burden requires 2 dimensionless parameters, instead of single *R*_0_

**Figure F:**
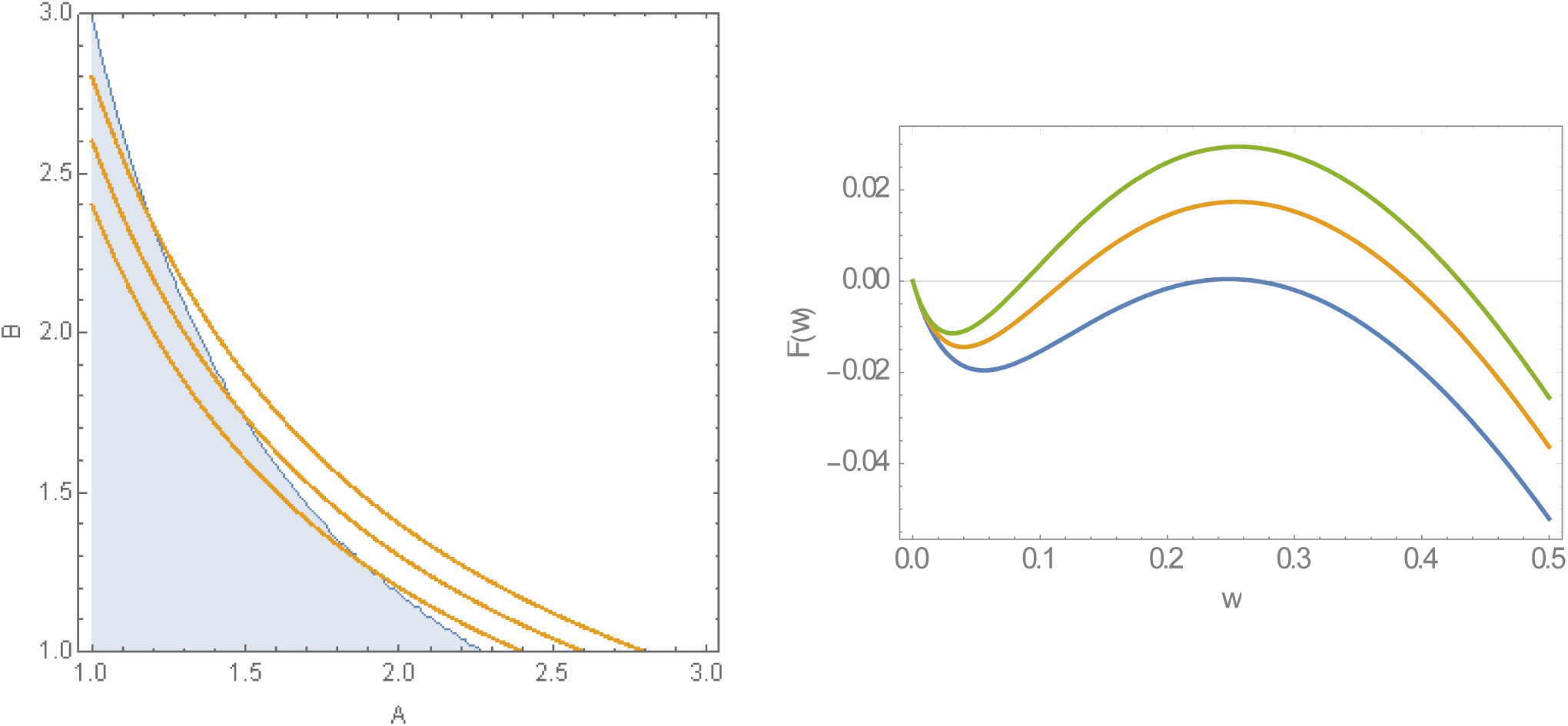
(a) (A, B) parameter space of MacDonald system with mating for rescaled parameters A → A / (γ + μ), B → B /ν. Shaded region is ‘stable zero’ (elimination case); open region has triple (bistable) equilibria. Orange lines are isocontours of R_0_ = AB = {2.4, 2.6, 2.8}. (b) Functions F (w | A, B) of reduced MacDonald system, for fixed R_0_ = 2.6, and 3 values R_0_ = 2.6; A = 1.5 is close to bifurcation (zero->triple equilibria). A = {1.5, 2, 2.5}. Case R_0_ = 2.6;*A* is close to bifurcation (zero->triple equilibria).

Figure F(a) shows that ‘endemic persistence - elimination’ regions in the (A,B) –parameter space– deviate markedly from isocontours of *R*_0_. This deviation renders it useless for analysis/prediction of long-term outcomes.

### Periodic MDA for MacDonald systems

Here we shall use stationary transmission environment, but vary transmission coefficients. Specifically, we fixed *R*_0_ = 3, and took 4 A-values in the range 2-3.5. Figure G (a) shows the resulting reduced MacDonald functions (Box 1), and equilibria (zero – breakpoint - endemic). As transmission coefficients are proportional to population densities (*A* ∝ *N* - snail, *B* ∝ *H* - human), different (snail per human) A/B ratio correspond to different transmission environments, and under identical *R*_0_ they can produce different outcomes, Figure G (b). In all 4 scenarios, disease is eliminated in finite time, once prevalence falls below the breakpoint. But higher A (large snail-to-human ratio) have stronger rebound and require more MDA rounds. Figure G (b) show MDA histories for the reduced MacDonald system.

Next we compared MDA predictions of the reduced model with the full host-vector MacDonald system, panels (c-f) of Figure G. The two models have comparable predictions at low transmission *A* = 2, but they diverge markedly as *A* increases. In most cases the reduced model exhibits stronger post-MDA rebound, and therefore takes longer to eliminate, or to reach a breakpoint. The extreme case is *A* = 3.5 (panel (f)), where complete model goes to elimination after 25 years of MDA, while the reduced model is locked in a limit cycle

**Figure G:**
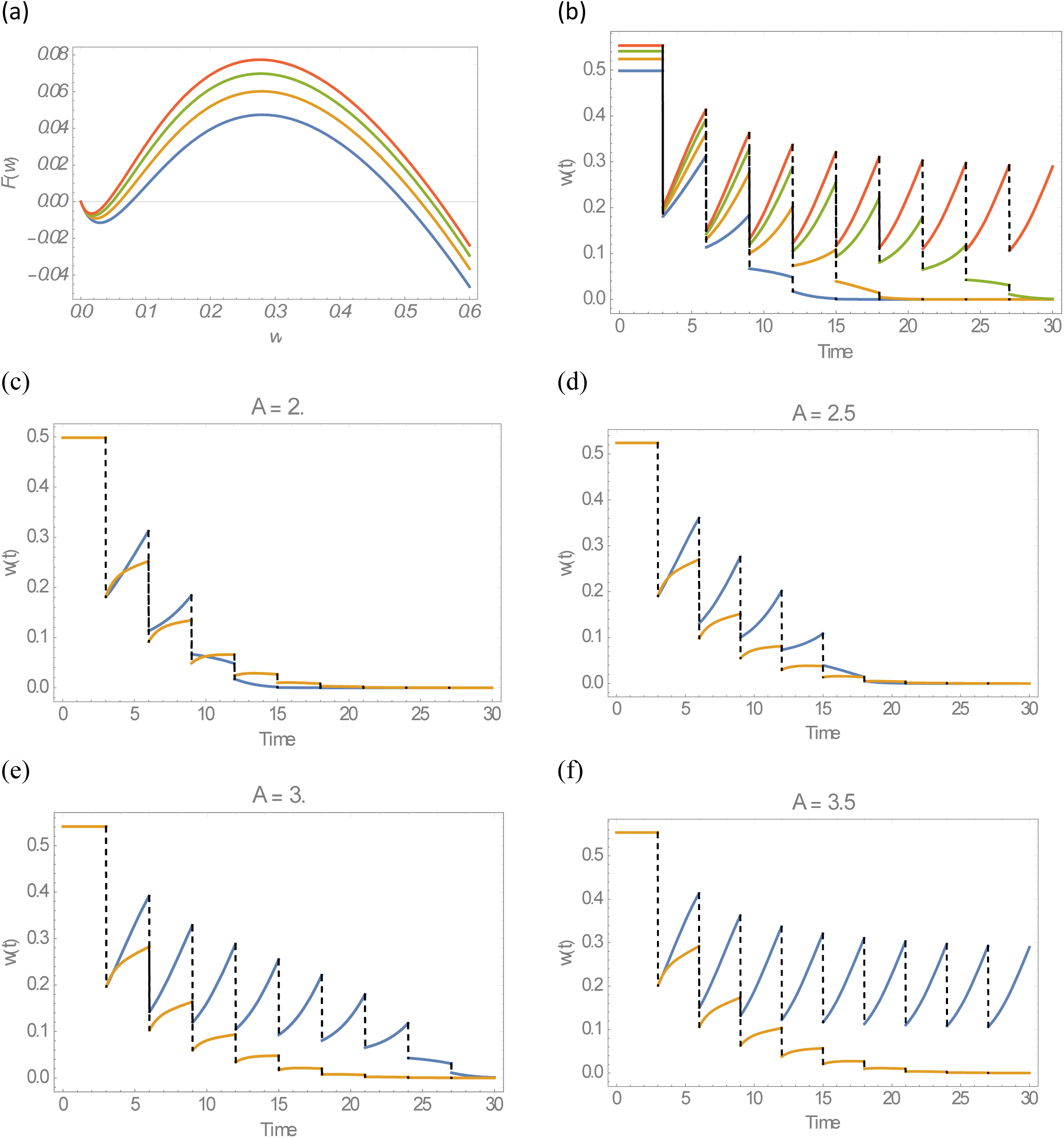
MDA control of MacDonald system with mating, fixed R_0_ = 3, and 4 vales of transmission coefficient A = 2 − 3.5. Panel (a) shows the corresponding reduced MacDonald functions **Error! Reference source not found**. and equilibria. Panel (b) MDA histories of 4 communities. Panels (c-f) compare MDA histories of the reduced model (blue) and the complete MacDonald (yellow), for 4 choices of A.

### Dynamic human-snail contacts in MacDonald system

Seasonality can affect both snail population biology, and human-snail contact patterns.

The corresponding MacDonald system (assuming stationary snail) turns into

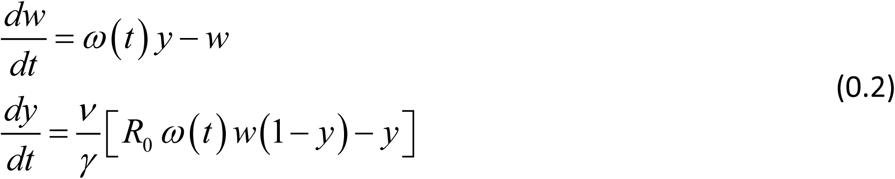

Its reduced form

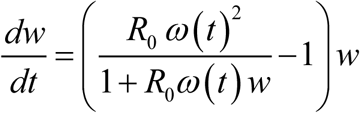

#### Dynamic snail population

Seasonal snail population in MacDonald system has significant effect on its dynamic patterns. Figures below illustrate for a range of *R*_0_ and variability parameter

**Figure H:**
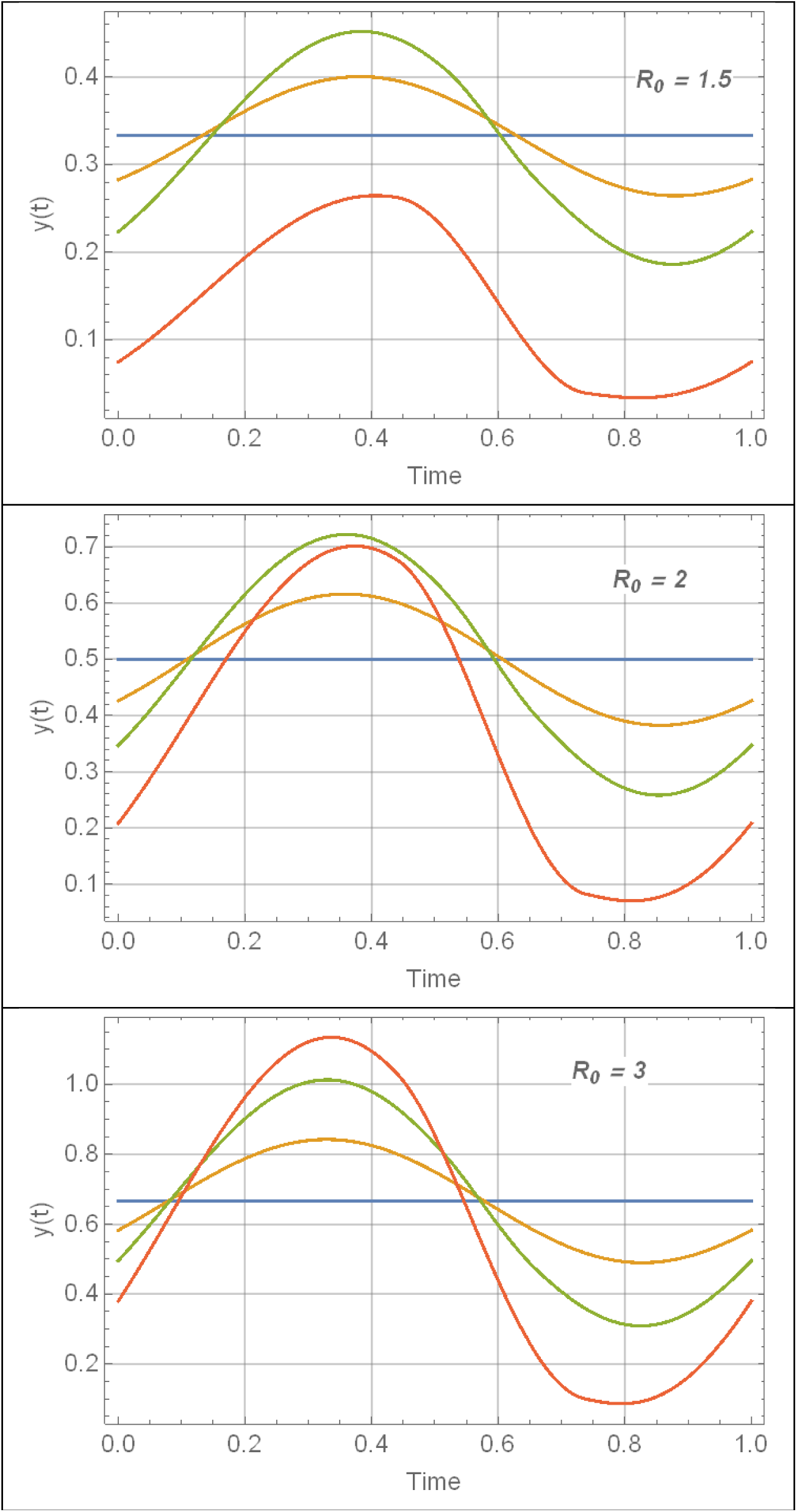
Periodic snail solutions y(t) for 3 R_0_ values of Table 1, and amplitude range 0 ≤ α ≤ .9 (blue to red).

**Figure I:**
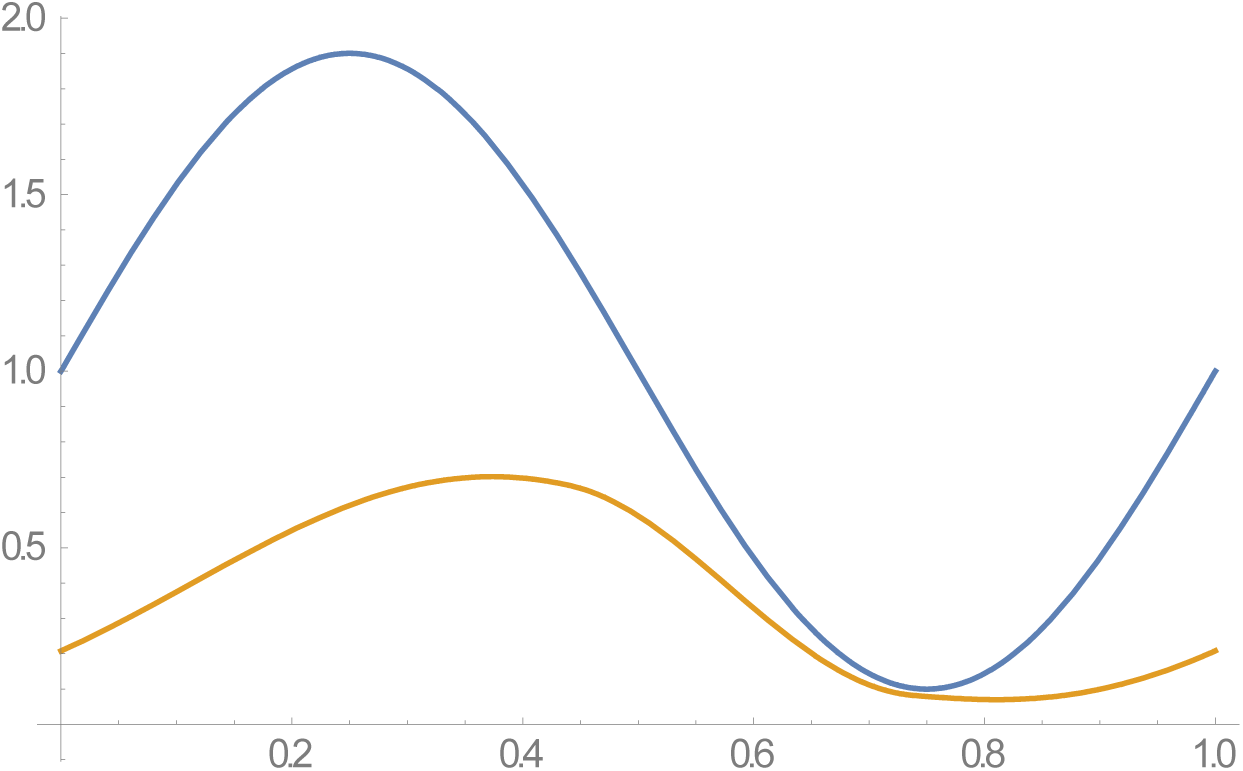
*Sustained seasonal patterns of the reduced MacDonald system and* complete host-vector systems for *R*_0_ = 2. Total snail *N* (*t*) (blue), infected *y* (*t*) (yellow). Rapid decline of N(t), brings y(t) close to N.

## References

1. Anderson RM, May RM. Infectious diseases of humans: dynamics and control. Oxford university press; 1992.

2. Anderson RM, May RM. Infectious Disease of Humans: Dynamics and Control. New York: Oxford University Press; 1991.

3. Anderson R, Turner H, Farrell S, Truscott J. Studies of the transmission dynamics, mathematical model development and the control of schistosome parasites by mass drug administration in human communities. Advances in parasitology. Vol 94: Elsevier; 2016:199–246.

4. Macdonald G. The dynamics of helminth infections, with special reference to schistosomes. Transactions of the Royal Society of Tropical Medicine and Hygiene. 1965;59(5):489–506.

5. Nasell I. Mating Models for Schistosomes. Journal of Mathematical Biology. 1978;6(1):21–35.

6. Barbour AD. Macdonalds Model and Transmission of Bilharzia. Transactions of the Royal Society of Tropical Medicine and Hygiene. 1978;72(1):6–15.

7. May RM. Togetherness among schistosomes: its effects on the dynamics of the infection. Mathematical biosciences. 1977;35(3):301–343.

8. Gurarie D, King CH, Yoon N, Li E. Refined stratified-worm-burden models that incorporate specific biological features of human and snail hosts provide better estimates of Schistosoma diagnosis, transmission, and control. Parasit Vectors. 2016;9(1):428.

9. Gurarie D, King CH. Population biology of Schistosoma mating, aggregation, and transmission breakpoints: more reliable model analysis for the end-game in communities at risk. PLoS One. 2014;9(12):e115875.

10. Gurarie D, King CH, Wang X. A new approach to modelling schistosomiasis transmission based on stratified worm burden. Parasitology. 2010;137(13):1951–1965.

11. Kariuki HC, Clennon JA, Brady MS, et al. Distribution patterns and cercarial shedding of Bulinus nasutus and other snails in the Msambweni area, Coast Province, Kenya. The American journal of tropical medicine and hygiene. 2004;70(4):449–456.

12. Ouma JH, Sturrock RF, Klumpp RK, Kariuki HC. A Comparative-Evaluation of Snail Sampling and Cercariometry to Detect Schistosoma-Mansoni Transmission in a Large-Scale, Longitudinal Field-Study in Machakos, Kenya. Parasitology. 1989;99:349–355.

13. Sturrock RF, Diaw OT, Talla I, Niang M, Piau JP, Capron A. Seasonality in the transmission of schistosomiasis and in populations of its snail intermediate hosts in and around a sugar irrigation scheme at Richard Toll, Senegal. Parasitology. 2001;123(SUPPL.):S77–S89.

14. Jordan P. Schistosomiasis: The St. Lucia Project. Cambridge: Cambridge University Press; 1985.

15. Remais J, Hubbard A, Wu ZS, Spear RC. Weather-driven dynamics of an intermediate host: mechanistic and statistical population modelling of Oncomelania hupensis. Journal of Applied Ecology. 2007;44(4):781–791.

16. Perez-Saez J, Mande T, Ceperley N, et al. Hydrology and density feedbacks control the ecology of intermediate hosts of schistosomiasis across habitats in seasonal climates. Proc Natl Acad Sci U S A. 2016;113(23):6427–6432.

17. Liang S, Seto EYW, Remais JV, et al. Environmental effects on parasitic disease transmission exemplified by schistosomiasis in western China. Proceedings of the National Academy of Sciences. 2007;104(17):7110–7115.

18. Mari L, Ciddio M, Casagrandi R, et al. Heterogeneity in schistosomiasis transmission dynamics. J Theor Biol. 2017;432:87–99.

19. Gurarie D, King C, Yoon N, Wang X, Alsallaq R. Seasonal dynamics of snail populations in coastal Kenya: Model calibration and snail control. Advances in Water Resources. 2017;108:397–405.

20. Nasell I. Hybrid Model of Schistosomiasis with Snail Latency. Theoretical Population Biology. 1976;10(1):47–69.

21. Anderson RM, May RM. Helminth Infections of Humans - Mathematical-Models, Population-Dynamics, and Control. Advances in Parasitology. 1985;24:1–101.

22. Rabone M, Wiethase JH, Allan F, et al. Freshwater snails of biomedical importance in the Niger River Valley: evidence of temporal and spatial patterns in abundance, distribution and infection with Schistosoma spp. Parasites & vectors. 2019;12(1):498.

23. Li E, Gurarie D, Lo NC, Zhu X, King CH. Improving public health control of schistosomiasis with a modified WHO strategy: a model-based comparison study. The Lancet Global Health. 2019;to appear.

24. Li EY, Gurarie D, Lo NC, Zhu X, King CH. Improving public health control of schistosomiasis with a modified WHO strategy: a model-based comparison study. The Lancet Global Health. 2019;7(10):e1414–e1422.

25. Lo NC, Gurarie D, Yoon N, et al. Impact and cost-effectiveness of snail control to achieve disease control targets for schistosomiasis. Proc Natl Acad Sci U S A. 2018;115(4):E584–E591.

26. Gurarie D, Lo NC, Ndeffo-Mbah ML, Durham DP, King CH. The human-snail transmission environment shapes long term schistosomiasis control outcomes: Implications for improving the accuracy of predictive modeling. PLoS Negl Trop Dis. 2018;12(5):e0006514.

27. King CH, Bertsch D. Historical perspective: snail control to prevent schistosomiasis. PLoS neglected tropical diseases. 2015;9(4):e0003657.

